# Climate change and environmental pollutants – an international survey of naturopathic perceptions and clinical behaviour

**DOI:** 10.64898/2026.05.31.26354564

**Authors:** Hope Foley, Iva Lloyd, Moira Fitzpatrick, Amie Steel

**Affiliations:** National Centre for Naturopathic Medicine, Faculty of Health, Southern Cross University, NSW, Australia; Australian Research Consortium in Complementary and Integrative Medicine, Faculty of Health, University of Technology Sydney, NSW, Australia; World Naturopathic Federation, Toronto, Ontario, Canada

**Keywords:** Planetary health, Environmental health, Climate change, Traditional, complementary and integrative medicine, Patient care, Health literacy, Naturopathy

## Abstract

**Background:** With rising concerns about health impacts from climate change and environmental exposures, planetary health approaches are increasingly prominent, considering connections between human health and that of the natural environment. Naturopathy is an holistic traditional medicine system characterised by philosophies and practices rooted in nature that theoretically align with planetary health. However, it is unknown to what extent these philosophies translate into consideration of relevant factors during patient care. This study describes the perceptions and clinical behaviours of the global naturopathic workforce in addressing the health impacts of climate change and environmental pollutants.

**Methods:** A cross-sectional online survey was administered to an international sample of naturopathic practitioners, recruited through communications from World Naturopathic Federation member organisations. The survey utilised the Climate Change Perceptions Scale, and asked participants about their perceptions of the health impacts of climate change and environmental pollutants. The survey also examined participant considerations of factors relating to climate change and environmental pollutants during clinical case assessment and prescribing of treatments. Data were descriptively analysed.

**Results:** Of n=363 naturopathic practitioners who completed the survey, 88.7% agreed climate change is real, of whom the majority were concerned about impacts of climate change on their patients’ health (89.1%). Almost all participants agreed that environmental pollutants harm human health (99.7%) and were concerned about impacts on their patients (99.5%). Climate-related health factors such as water intake (74.2%) and food security (72.9%) were frequently considered during patient assessment, while impacts of severe weather events (41.4%) were less commonly considered. Consideration of factors relating to environmental pollutants was more commonly reported, particularly for food quality (83.8%) and domestic/indoor sources of pollutants (73%). When formulating prescriptions, participants reported highly frequent consideration of all climate-related factors (73%-86.8%) and varied consideration of environmental pollutant exposures (54.4%-83.4%).

**Conclusions:** The global naturopathic workforce demonstrates a high level of awareness and engagement with factors relating to health impacts of climate change and environmental pollutants, suggesting alignment with planetary health. While this engagement is evident in clinical behaviour, some gaps between awareness and application suggest a need for greater support to strengthen the naturopathic application of planetary and environmental health.

## Introduction/background

Naturopathy is a system of traditional medicine that originated in Europe and is now practiced in over 108 countries, spanning all World Health Organization (WHO) World Regions [1]. The World Naturopathic Federation (WNF) – the international representative body for the naturopathic profession [2] – defines naturopathy according to its traditional philosophies and principles [3]. Naturopathic principles include: *First Do No Harm, Doctor as Teacher, the Healing Power of Nature, Treat the Whole Person, Treat the Cause, Wellness, Health Promotion and Disease Prevention* [1]. The naturopathic principles are applied through detailed, individualised patient assessment that considers: diet and lifestyle behaviours, personal and family health history, social determinants of health, and environmental factors [1,3]. This extends to assessment of pollutants and other environmental exposures as potential causal factors to symptoms or health conditions [3], which can align naturopathy with planetary health approaches.

Planetary health addresses the inextricable connection between the health of human populations and that of the natural systems on which human life depends [4]. Planetary health considers factors such as climate change - the long-term changes in the climatological and weather patterns that had come to define Earth’s local, regional and global climates, primarily caused by human-made emissions of carbon and other greenhouse gases [5]. Climate change impacts health both directly and indirectly; extreme heat and weather events present direct risks, while also leading to the disruption of safe food and water access, displacement of communities, and increases in zoonoses and vector-borne diseases [6].

Planetary health also promotes public health approaches inclusive of environmental health, which addresses environmental exposures such as poor air and water quality or toxic pollutants [7]. Environmental pollutants contaminate the environment, originating from sources such as air pollution, industrial discharges, and the accumulation of toxic chemicals in water and soil, including organic compounds and heavy metals [8]. Environmental pollutants are known to have profound, broad-ranging impacts on health [8]. Air pollution alone causes approximately 7 million deaths each year globally[9] and is linked to cognitive decline and neurodegenerative diseases [10]. Other pollutants can include endocrine disrupting compounds associated with cancer, cardiovascular problems, and reproductive concerns [11], or microplastics associated with immunological, respiratory, cardiovascular, gastrointestinal and endocrine dysfunction [12]. The consideration of environmental exposures (environmental pollutants, climate factors) as causal contributors to symptoms and health conditions is commonly included in naturopathic assessment [13], yet appears to be lacking in routine biomedical assessment [14], despite established identification of these exposures as health risks by the WHO [6,7,15], and a growing research base [9,16].

Public concern about climate change, environmental toxins, and their health impacts has increased substantially over the past two decades. For example, Canadian population surveys indicate concern about climate change rose from 69% in 2006 to 79% in 2017 and 85% in 2022 [17]. Similar rates of concern or increases in public engagement with the issue of climate change and health have been found in recent studies from Germany [18], Egypt [19], Bangladesh [20], and Australia [21], as well as international studies [22,23]. However, research also indicates that public perceptions of climate change and environmental toxins are shaped by complex factors, including political orientation [24], governmental and media focus [23], and inconsistent terminology (i.e., interchangeable use of terms such as *greenhouse effect*, *global warming*, and *climate change*, which can complicate interpretation and communication) [25]. Similarly, concern about climate change amongst healthcare professionals has garnered increasing focus in recent years as healthcare systems and providers increasingly experience the impacts of climate change on patient health and care provision [26]. Subsequently, there is growing interest in the sustainability of healthcare and health systems’ responsiveness to climate-related impacts, including practitioner knowledge, literacy and capacity to take relevant actions [26–29].

The naturopathic profession demonstrates active interest in environmental and planetary health, including the development of dedicated groups [30], calls to action [31], and contributions to identifying links between environmental pollutants and key non-communicable diseases [32]. The WNF also offers detailed public resources on environmental health and climate change on a dedicated website [33], and research has begun to explore the role of naturopathic practitioners during climate disasters [34]. Despite this engagement from the profession, there is very little research exploring the perceptions of naturopathic practitioners with respect to environmental pollutants and climate change, and the respective role of naturopathic care. This study aims to describe the perceptions and clinical behaviours of the global naturopathic workforce in addressing the health impacts of climate change and environmental pollutants.

## Methods

### Study design

This study employed a cross-sectional survey design, administered online with an international sample. The study conduct and reporting were aligned with the Strengthening the Reporting of Observational Studies in Epidemiology (STROBE) Statement [35], including items specific to cross-sectional studies (completed checklist provided in Additional File 1).

### Participants, sampling and recruitment

Participants were naturopathic practitioners from any country who reported being active in clinical practice for at least five years and were able to complete the survey in one of the available languages. A sample size of approximately 385 was sought based on standard calculations for descriptive survey research and pilot studies [36]. Due to the descriptive nature and novelty of the research aim, this was considered sufficient to provide a meaningful descriptive analysis.

The WNF facilitated recruitment with the assistance of their member organisations. The WNF emailed information about the study with a link to the online survey to each of their full member organisations, representing national naturopathic associations from 33 countries. Each member organisation was asked to share the information and survey link with their members via email and social media. The survey information and link were also shared on the WNF social media pages. Naturopathic practitioners were then able to opt in for participation and proceed with the survey after reading the information and providing consent. Eligibility criteria were checked during the provision of consent. The survey information, recruitment materials and online survey were all provided in four languages (English, French, Spanish, Portuguese), selected on the basis of previous global naturopathic studies to optimise accessibility [37,38].

### Instrument and data collection

The survey was hosted on the Qualtrics™ survey administration platform, accessed via an anonymous link, and took approximately 12-18 minutes. Participants self-selected their preferred language. The survey instrument and information sheet were developed in English and translated with the support of WNF volunteer translators who were naturopathic professionals fluent in their respective language as well as English. The translation process initially utilised the Qualtrics auto-translate function to produce draft documents in Spanish, Portuguese and French. The relevant translated draft was provided to each translator alongside the original English copy, and the translators edited the translations for accuracy in meaning and context.

The instrument comprised 65 items across five sections: 1) Perceptions of climate change and environmental health, 2) Climate and environmental health in naturopathic case assessment, 3) Naturopathic treatments and care for the health impacts of climate change and environmental toxins, 4) Advocacy for climate and environmental health, and 5) Participant demographics and practice characteristics. Analyses presented in this paper draw from sections 1, 2, 3 and 5, as outlined below. The full survey is provided in Additional File 2.

#### Perceptions of climate change and environmental health

The survey began with the Climate Change Perceptions Scale (CCPS), a validated measure of five items beginning with statement “I believe that climate change is real”, followed by four items about the causes and consequences of climate change [39]. Participants who responded ‘Strongly disagree’ or ‘disagree’ to the first item were not shown the remaining four items. An additional health-specific set of ten items were developed for this section to allow participants to rate the degree to which they were concerned about the impacts of climate change (6 items) and environmental pollutants (4 items) on the health of their patients and broader community, as well as 3 items about concerns they had encountered from patients on the same topics. All items in this section employed a five-point scale from ‘Strongly agree’ to ‘Strongly disagree’, except the 3 items on patient concerns which were rated from ‘Always’ to ‘Never’.

#### Climate and environmental health in naturopathic case assessment

The second section of the survey asked participants about observations and behaviours they undertake during clinical case assessment regarding health issues related to climate change (6 items) and environmental health (8 items). These items were drafted to capture information about relevant priority issues highlighted in international public health guidance about climate and health [40], framed within the context of naturopathic practice [3]. This included factors such as food security and quality, water availability and quality, air quality, extreme weather events, and related health outcomes. Participants rated how frequently they consider each factor in patient case assessment using a five-point scale from ‘Always’ to ‘Never’.

#### Naturopathic treatments and care for the health impacts of climate change and environmental toxins

The survey included questions about the treatments, prescriptions and advice participants provide during naturopathic care regarding factors related to climate change (6 items) and environmental health (7 items). These items aligned with the same priority health issues covered in the previous section, allowing participants to rate how frequently they consider each factor when applying or selecting naturopathic interventions and advice from ‘Always’ to ‘Never’.

#### Participant demographics and practice characteristics

The final survey section captured participant demographics, alongside professional and practice characteristics, using multiple choice and open-text items. Three demographic items recorded participant location (country of residence), gender and age. Practice characteristics were reported across five items including location (country of clinical practice, if different from residence), years spent in clinical practice, clinical specialisation/special focus, clinical setting, and clinic locale (urban, regional, rural and/or online).

### Data handling and analysis

Data were exported from Qualtrics into StataSE 18 for cleaning and analysis. Observations were considered incomplete and removed from the dataset if more than 20% of responses were missing. To combat the data integrity challenges of online survey research, the Qualtrics embedded reCAPTCHA function scores were utilised alongside standard data integrity checks for straight-lining, very low completion time, inconsistent responses and nonsensical open-text responses to remove observations indicating a high likelihood of bots or disengagement. A descriptive analysis was undertaken, reporting data as frequencies and percentages. Scale categories for variables relating to perceptions of climate change and environmental toxins were collapsed to reflect agreement (agree/strongly agree), disagreement (disagree/strongly disagree) or neutrality (neither agree nor disagree). Similar re-categorisation was undertaken with variables regarding clinical considerations to reflect routine consideration (always/most of the time), some consideration (sometimes), or infrequent consideration (rarely/never). Missing data were excluded and indicated via reported n values.

### Ethical considerations

The study protocol and ethical considerations were drafted in accordance with the Declaration of Helsinki. Ethical approval was sought and obtained from the Human Research Ethics Committee at the University of Technology Sydney (ETH24-10002).

## Results

During data cleaning, 71 responses were removed due to data integrity checks indicating a high likelihood of being generated by bots, and 81 were removed as incomplete. A total of n=363 complete responses comprised the final sample for analysis. As it was not possible to know how many potential participants received an invitation or saw a social media post about the study, a participation rate of 57% was calculated (following the removal of responses that failed data integrity checks) based on the number of complete responses (n=363) divided by the total number of people who clicked through to access the participant information sheet (n=640).

### Participant and practice characteristics

Participant and practice characteristics are shown in Table 1. The final sample included participants from five of the six WHO World Regions, with the highest representation coming from the Region of the Americas (n=191, 53.3%) and the lowest from the African Region (n=11, 3.1%), while the Eastern Mediterranean Region was not represented. Participants were based in 32 countries, predominantly the USA (n=99, 27.7%), France (n=66, 18.4%) and Canada (n=45, 12.6%). All four language versions of the survey were utilised, with the English language copy being accessed the most (n=248, 68.3%) and the Spanish copy accessed the least (n=22, 6.1%). Participants predominantly identified as female (n=272, 75.6%) and were typically aged between 45-54 years (n=107, 30.4%) or 35-44 years (n=92, 26.1%).

**Table 1.** Participant demographics and practice characteristics.

|  | n | % |
| --- | --- | --- |
| **Participating naturopathic practitioners** | 363 | 100 |
| **WHO World Region** |  |  |
| Region of the Americas – North America | 144 | 40.2 |
| Region of the Americas – Latin America | 47 | 13.1 |
| European Region | 89 | 24.9 |
| Western Pacific Region | 40 | 11.2 |
| South-East Asian Region | 27 | 7.5 |
| African World Region | 11 | 3.1 |
| **Survey language selected** |  |  |
| English | 248 | 68.3 |
| French | 65 | 17.9 |
| Portuguese | 28 | 7.7 |
| Spanish | 22 | 6.1 |
| **Gender** |  |  |
| Female | 272 | 75.6 |
| Male | 86 | 23.9 |
| Non-binary | 2 | 0.6 |
| **Age** |  |  |
| 24-34 years | 58 | 16.5 |
| 35-44 years | 92 | 26.1 |
| 45-54 years | 107 | 30.4 |
| 55-64 years | 65 | 18.5 |
| 65 years and over | 30 | 8.5 |
| **Years in naturopathic practice** |  |  |
| Up to 10 years | 161 | 44.5 |
| 11-15 years | 70 | 19.3 |
| 16-20 years | 54 | 14.9 |
| 21-25 years | 30 | 8.3 |
| More than 25 years | 47 | 13.0 |
| **Specialisation in practice** |  |  |
| Generalist practice | 200 | 55.1 |
| Environmental health specialisation | 42 | 11.6 |
| Climate and health specialisation | 10 | 2.8 |
| Other area of specialisation | 128 | 35.3 |
| **Clinic environment. I am in a clinic...** |  |  |
| By myself | 134 | 36.9 |
| With other health professionals but no other naturopathic practitioners | 77 | 21.2 |
| With other naturopathic practitioners and other health professionals | 77 | 21.2 |
| With other naturopathic practitioners but no other types of health professionals | 27 | 7.4 |
| Other setting | 31 | 8.5 |
| I am in a hospital setting | 17 | 4.7 |
| **Clinic locality** |  |  |
| Urban or suburban | 236 | 65.2 |
| Regional area | 63 | 17.4 |
| Rural or remote | 58 | 16.0 |
| Online | 47 | 13.0 |

Less than half of participants were in their first decade of clinical practice (n=161, 44.5%), while n=47 (13%) had been in practice for over 25 years. Over half of participants (n=200, 55.1%) reported taking a generalist approach to practice, while others reported one or more areas of specialisation, including environmental health (n=42, 11.6%), climate-related health (n=10, 2.8%) and/or other areas of specialisation (n=128, 35.3%). Participants were most commonly in solo clinical practice (n=134, 36.9%), or sharing with either a mix of other naturopathic and non-naturopathic health care providers (n=77, 21.2%) or only non-naturopathic providers (n=77, 21.2%). Sharing exclusively with other naturopathic practitioners was less common (n=27, 7.4%) and hospital settings were reported the least (n=17, 4.7%). Almost two-thirds of participants reported practicing from an urban or suburban locality (n=236, 65.2%) while online settings were the least-common practice location (n=47, 13.0%).

### Perceptions of climate change, environmental toxins and related health impacts

Participant perceptions of climate change and related health impacts are detailed in Table 2. The base item for the CCPS revealed the vast majority of participants (n=322, 88.7%) strongly agreed or agreed they believe climate change is real, and a small number neither agreed nor disagreed (n=17, 4.7%). The remaining n=24 (6.6%) participants who strongly disagreed or disagreed that climate change is real skipped the four remaining items in the CCPS and the four items about health impacts related to climate change.

**Table 2.** Perceptions of climate change, health impacts of climate change and environmental pollutants, and associated patient concerns.

| Item | Strongly agree or agree |  | Neither agree nor disagree |  | Disagree or Strongly disagree |  |
| --- | --- | --- | --- | --- | --- | --- |
|  | n | % | n | % | n | % |
| **Base item (n=363)** |  |  |  |  |  |  |
| I believe that climate change is real | 322 | 88.7 | 17 | 4.7 | 24 | 6.6 |
| **Climate change perception scale (n=339)\*** |  |  |  |  |  |  |
| Climate change will bring serious negative consequences | 311 | 91.7 | 25 | 7.4 | 3 | 0.9 |
| My local area will be influenced by climate change | 308 | 90.9 | 26 | 7.7 | 5 | 1.5 |
| The main causes of climate change are human activities | 293 | 86.4 | 35 | 10.3 | 11 | 3.2 |
| It will be a long time before the consequences of climate change are felt | 58 | 17.1 | 49 | 14.5 | 232 | 68.4 |
| **Perceptions of health impact of climate change (n=339)\*** |  |  |  |  |  |  |
| Climate change can affect human health | 324 | 95.6 | 13 | 3.8 | 2 | 0.6 |
| Climate change is already affecting human health | 315 | 92.9 | 21 | 6.2 | 3 | 0.9 |
| I am concerned about the potential impacts of climate change on human health | 309 | 91.2 | 23 | 6.8 | 7 | 2.1 |
| I am concerned about the potential impacts of climate change on the health of my patients | 302 | 89.1 | 24 | 7.1 | 13 | 3.8 |
| **Perceptions of health impact of environmental pollutant (n=363)** |  |  |  |  |  |  |
| Environmental pollutants such as those found in the air, water, food or household environments are having negative impacts on human health | 362 | 99.7 | 0 | 0.0 | 1 | 0.3 |
| I am concerned about the effects of environmental pollutants on human health | 360 | 99.2 | 3 | 0.8 | 0 | 0.0 |
| I am concerned about the effects of environmental pollutants on the health of my patients<sup>†</sup> | 360 | 99.5 | 2 | 0.6 | 0 | 0.0 |
|  | **Always or Most of the time** |  | **Sometimes** |  | **Rarely or never** |  |
| **How often do your patients express the following concerns about climate and environmental health? (n=362)** | n | % | n | % | n | % |
| My patients express concerns about climate change impacting their physical health | 83 | 22.9 | 151 | 41.7 | 128 | 35.4 |
| My patients express concerns about climate change impacting their mental health | 74 | 20.4 | 138 | 38.1 | 150 | 41.4 |
| My patients express concerns about environmental pollutants or other environmental factors impacting their health | 161 | 44.5 | 164 | 45.3 | 37 | 10.2 |

\*Items not displayed to participants who responded 'Disagree' or 'Strongly disagree' to Base item.
<sup>†</sup>One missing value

Of the 339 (93.4%) participants who completed all CCPS items, most strongly agreed or agreed: the main causes of climate change are human activities (n=293, 86.4%), climate change will bring serious negative consequences (n=311, 91.7%), and their local area will be influenced by climate change. More than two-thirds of respondents (n=232, 68.4%) disagreed or strongly disagreed that it will be a long time before the consequences of climate change are felt. Regarding the health-related impacts of climate change, the vast majority of respondents strongly agreed or agreed that climate change can affect human health (n=324, 95.6%) and is already affecting human health (n=315, 92.9%). It was also typical for respondents to strongly agree or agree they are concerned about the potential impacts of climate change on human health (n=309, 91.2%) and the health of their patients (n=302, 89.1%). Participants commonly reported their patients expressed concerns about climate change impacting on their physical health either sometimes (n=151, 41.7%) or always/most of the time (n=83, 22.9%), while the remaining 35.4% (n=128) heard this concern rarely or never. Encountering patient concerns about the mental health impacts of climate change was reported at a slightly lower frequency.

Almost all participants strongly agreed or agreed (n=362, 99.7%) that environmental pollutants are having negative impacts on human health, with only one (0.3%) participant in disagreement. Similarly, participants almost exclusively agreed or strongly agreed they were concerned about the effects of environmental pollutants on human health (n=360, 99.2%) and the health of their patients (n=360, 99.5%). The vast majority of participants reported they had heard patients express concerns about the health impacts of environmental pollutants and other environmental factors either sometimes (n=164, 45.3%) or always/most of the time (n=161, 44.5%), while only 10.2% (n=37) of participants reported never or rarely hearing this patient concern. Full details are shown in Table 2.

### Consideration of climate and environmental factors during clinical case assessment

Table 3 shows participant responses regarding how often they consider various climate-related and environmental factors during the clinical assessment of patient cases. The most commonly reported climate-related factors that participants considered always or most of the time were sufficient water intake for the patient’s local climate and health status (n=268, 74.2%) and the patient’s ability to access fresh, healthy food (n=264, 72.9). The least considered climate factor was the impact of weather changes or severe weather events on the patient’s health and wellbeing, which was considered always or most of the time by 41.4% (n=150) of participants, with almost one-quarter (n=84, 23.2%) reporting they rarely or never consider this factor.

**Table 3.** Reported consideration of climate-related and environmental factors during <u>clinical assessment of patient case</u>.

| Item | Total responses | Always or Most of the time |  | Sometimes |  | Rarely or never |  |
| --- | --- | --- | --- | --- | --- | --- | --- |
|  | n | n | % | n | % | n | % |
| **Consideration of CLIMATE-RELATED factors during case assessment** |  |  |  |  |  |  |  |
| Sufficient water intake for the patient's local climate and health status | 361 | 268 | 74.2 | 51 | 14.1 | 42 | 11.6 |
| The patient's ability to access fresh, healthy food | 362 | 264 | 72.9 | 54 | 14.9 | 44 | 12.2 |
| The patient's ability to access clean drinking water | 363 | 237 | 65.3 | 48 | 13.2 | 78 | 21.5 |
| The environmental sustainability of patients' dietary habits | 362 | 211 | 58.3 | 96 | 26.5 | 55 | 15.2 |
| Patient's exposure to external air pollution from climate change or extreme weather events | 363 | 204 | 56.2 | 102 | 28.1 | 57 | 15.7 |
| Impact of weather changes or severe weather events on the patient's health and wellbeing | 362 | 150 | 41.4 | 128 | 35.4 | 84 | 23.2 |
| **Consideration of ENVIRONMENTAL factors during case assessment** |  |  |  |  |  |  |  |
| External environmental factors affecting food quality for patients | 363 | 304 | 83.8 | 50 | 13.8 | 9 | 2.5 |
| Patient's exposure to domestic/indoor sources of air pollution in living/working environments | 363 | 265 | 73.0 | 88 | 24.2 | 10 | 2.8 |
| Patient's exposure to external sources of air pollution in living/working environments | 363 | 261 | 71.9 | 80 | 22.0 | 22 | 6.1 |
| Domestic environmental factors affecting food quality for patients | 362 | 259 | 71.6 | 83 | 22.9 | 20 | 5.5 |
| Patient's exposure to other environmental toxins in the home, work or natural environments | 361 | 242 | 67.0 | 92 | 25.5 | 27 | 7.5 |
| Domestic environmental factors affecting patient's water quality | 363 | 239 | 65.8 | 91 | 25.1 | 33 | 9.1 |
| External environmental factors affecting patient's water quality | 363 | 224 | 61.7 | 94 | 25.9 | 45 | 12.4 |
| Testing patient for presence of environmental toxic pollutants or associated impacts on health | 362 | 108 | 29.8 | 114 | 31.5 | 140 | 38.7 |

Environmental factors were reportedly considered more often than climate-related factors overall. External environmental factors affecting food quality for patients was the most commonly reported environmental factor category to be considered always or most of the time (n=304, 83.8%), followed by patients’ exposure to domestic/indoor sources of pollution in living/working environments (n=265, 73%). The vast majority of participants reported considering each environmental factor at least sometimes, with the exception of testing patients for the presence of environmental toxic pollutants or associated impacts on health, which 38.7% (n=140) of respondents reporting considering rarely or never.

### Consideration of climate-related factors and environmental toxins when formulating prescriptions and advice

The climate and environmental factors considered by participants during formulation of prescriptions and clinical advice are shown in Table 4. Climate-related factors were considered always or most of the time by a large majority of participants across all items, with the most commonly reported factors being the quality of food being recommended to or consumed by patients (n=315, 86.8%) and increasing time that patients spend in nature (nature prescribing) (n=314, 86.5%). The least commonly reported climate-related factors were consideration of environmental sustainability when prescribing herbs, supplements and other prescribed remedies (n=268, 73.8%) or dietary and food-based recommendations (n=275, 75.8%).

**Table 4.** Consideration of climate-related factors and environmental exposures when making recommendations or prescribing to patients.

| Item | Total<br>responses | Always or Most<br>of the time |  | Sometimes |  | Rarely or never |  |
| --- | --- | --- | --- | --- | --- | --- | --- |
| Consideration of CLIMATE-RELATED factors when prescribing | n | n | % | n | % | n | % |
| The quality of food being recommended to or consumed by patients | 363 | 315 | 86.8 | 52 | 11.6 | 6 | 1.7 |
| Increasing time that patients spend in nature (nature prescribing) | 363 | 314 | 86.5 | 41 | 11.3 | 8 | 2.2 |
| Educating patients about connection between human health and the natural environment | 362 | 306 | 84.5 | 45 | 12.4 | 11 | 3.0 |
| Educating patients about the health impacts of environmental pollutants | 363 | 306 | 84.3 | 50 | 13.8 | 7 | 1.9 |
| Environmentally sustainable dietary and food-based recommendations | 363 | 275 | 75.8 | 66 | 18.2 | 22 | 6.1 |
| Environmentally sustainable herbs, supplements and other prescribed remedies | 363 | 268 | 73.8 | 62 | 17.1 | 33 | 9.1 |
| Consideration of reducing exposure to ENVIRONMENTAL TOXINS in patients'... |  |  |  |  |  |  |  |
| Food | 361 | 301 | 83.4 | 44 | 12.2 | 16 | 4.4 |
| Water | 362 | 289 | 79.8 | 48 | 13.3 | 25 | 6.9 |
| Personal care products | 362 | 275 | 76.0 | 67 | 18.5 | 20 | 5.5 |
| Air | 361 | 249 | 69.0 | 76 | 21.1 | 36 | 10.0 |
| Home-based products used indoors | 362 | 244 | 67.4 | 91 | 25.1 | 27 | 7.5 |
| Home-based products used outdoors | 362 | 215 | 59.4 | 90 | 24.9 | 57 | 15.8 |
| Workplace products, tools and environments | 360 | 196 | 54.4 | 101 | 28.1 | 63 | 17.5 |

When asked how frequently they consider reducing exposure to environmental toxins during formulation of prescriptions and advice, most participants reported considering each factor at least sometimes, however, responses varied across the different types of exposure. The most commonly reported exposures that participants considered reducing for patients always or most of the time were in food (n=301, 83.4%), water (n=289, 79.8%) and personal care products (n=275, 76%). Workplace products, tools and environments (n=196, 54.4%), alongside home-based products used outdoors (n=215, 59.4%), were the exposure sources considered least.

## Discussion

This study provides the first global description of naturopathic practitioners’ perceptions of climate change and environmental pollutants, and how these issues are incorporated into the clinical practice context. Overall, the findings demonstrate a high level of awareness and concern among the naturopathic workforce regarding the health impacts of both climate change and environmental pollutants, with relevant considerations applied to practice. This highlights the current and potential capacity for naturopathic approaches to contribute meaningfully to environmental and planetary health. However, the results also reveal important gaps between professional awareness and consistent integration of these factors into routine patient assessment and care, highlighting areas for further education, resource development, and structural support.

The naturopathic workforce in this study expressed strong beliefs that climate change is real and serious, that both climate change and environmental pollutants can adversely affect human health, and that these factors should be considered in clinical practice. These findings support the view that naturopathic values and training align with environmental and planetary health considerations [41,42]. The rates of climate change belief and concern reported elsewhere by healthcare providers more broadly differ substantially across studies, making it difficult to draw direct comparisons between the naturopathic professionals in our sample and healthcare providers from other professions. The rates reported by our participants were similar to averages previously reported amongst environmental health professionals[43] and broad global samples of various healthcare professions [29,44], while being similar to or higher than rates reported in some studies with more localised samples [45]. Climate knowledge and concern amongst healthcare professionals – as well as preparedness to address related health impacts – may differ across factors such as profession [28], engagement with health politics [29], and personal or patient experiences of climate change impacts [27]. The nuance in the international landscape of climate change perceptions amongst healthcare providers suggests a need for education, training, resourcing and other support to equip the healthcare workforce in responding to climate-related health risks [45]. Additionally, while most of our participants acknowledged that climate change is primarily driven by human activity, an even greater proportion believed it will result in serious negative consequences. This divergence suggests concern about health impacts exist even among practitioners who hold varying views on causality, reinforcing the importance of clinical communication about climate that focus on health impacts and outcomes rather than on attribution of cause.

Interestingly, a greater proportion of participants reported concern about the impacts of environmental pollutants on human health than the impacts of climate change. A similar pattern emerged when considering impacts of these two exposure categories on patients’ health, and in regard to hearing patients express their own concerns. As naturopathic approaches have traditionally considered toxic exposures during patient assessment [3], this discrepancy may reflect a longer history of attention to the health risks of environmental pollutants, as well as influence from media reports about such pollutants [46,47] and routine labelling of risk-associated ingredients in food, personal care products and everyday items [48]. Environmental pollutants such as air pollution, chemical contaminants, and endocrine-disrupting compounds are directly associated with well-documented morbidity and mortality outcomes [9]. In contrast, climate change often affects health indirectly and over longer timeframes, which may reduce its perceived relevance and immediacy for both naturopathic practitioners in day-to-day practice [49] and for patients when considering their own health or healthcare [18]. Despite this, concerns about climate change and associated health impacts are prevalent in the general population [17,18,50], suggesting a need for further research into how climate and environmental issues are introduced, discussed, and understood by both parties in clinical encounters.

Environmental health factors were reported by many participants as routine considerations in naturopathic clinical assessment and care, which is consistent with extant naturopathic literature [1,3]. That is, assessing for environmental exposures such as the quality of air, water, and soil; environmental chemicals, heavy metals, and other pollutants; pathogens such as viruses, bacteria, mould, or fungi; and/or inquiring about the time spent in nature and exposure to sunlight [1,3]. There appears to be a gap between the awareness of the health impacts of environmental pollutants and climate change, and translation of that awareness to naturopathic clinical assessment. For example, consideration of specific environmental factors during clinical assessment (e.g., food quality and access, water access, exposure to air pollution and other environmental toxins) were reported at a much lower proportion than the more general awareness of environmental pollutants impacting human health. Additionally, when considering reduction of patient exposures to environmental toxins, a lower proportion of participants considered sources other than food and water. Specifically, less commonly reported exposures were the workplace, home, air, and personal care products, despite a growing evidence base linking exposures from these sources to health conditions [8,51] For example, air-borne pollutants found in homes and workplaces, such as particulate matter, tobacco smoke, asbestos, silica, and chemical products used in various industries from healthcare to hairdressing, have been linked to chronic respiratory diseases [52,53], while some gardening products may be carcinogenic [54] and personal care products often contain ingredients linked to inflammation, skin conditions [51] and endocrine disruption [55]. This gap between awareness and assessment may reflect multiple intersecting factors, including variability in educational preparation, time constraints during consultations, scope-of-practice limitations, and the absence of standardised environmental health assessment tools tailored to naturopathic practice [56,57]. It may also reflect the challenge of translating complex and systemic environmental risks into individual-level clinical assessments. The results indicate a need for further education and/or resources that naturopathic professionals can use to assist in patient education – a need reflected in some initiatives emerging from the global profession, such as the WNF Environmental Health website [33].

The findings of this study highlight existing strengths in naturopathic practice that provide a foundation for planetary and environmental health application. For example, food quality and access were indicated as primary factors included in both clinical assessment and prescribing of treatments, with food also being the primary source considered to reduce environmental pollutant exposures. This complements results from previous global surveys demonstrating the routine use of dietary prescriptions in naturopathic practice [38,58]. However, further research is needed to understand any amendments naturopathic practitioners are making to their dietary prescriptions in response to climate change and environmental exposures. Most respondents also reported recommending patients increase their amount of time spent in nature (nature prescribing), which reflects the naturopathic principle denoting the healing power of nature [3]. Nature prescribing is a field of social prescribing focused on engagement with natural environments as a health promoting behaviour and has drawn much recognition for its growing evidence-base in recent years [59]. However, the typically outdoor setting of nature prescribing is increasingly challenged by climate change and environmental pollution [60]. Ongoing research and practical action are required to provide guidance for healthcare providers to ensure nature prescriptions optimise benefits while minimising risks from weather events and environmental exposures. The key to success in this area may lie in the bidirectional relationship between human health and that of the natural environment, as knowledge of and engagement with the natural environment have been associated with pro-environmental attitudes and behaviours [61–63]. Given a high proportion of our respondents indicated that they educate patients about the connection between human and environmental health, naturopathic practitioners may be well-positioned to leverage this bi-directional relationship in practice [64].

While it was common for our participants to consider climate- and environment-related factors that affect patient health, rates of consideration were notably lower regarding prescribing behaviours that consider the environmental sustainability of practices or treatments used. The reduced incidence of these sustainability-related considerations - albeit still reported by more than 1 in 4 respondents - may reflect uncertainty about the extent to which individual clinical actions can meaningfully influence broader sustainability challenges [65], as well as limited training or resources addressing sustainable practice within clinical decision-making. Within the context of climate change and biodiversity loss, there has been much global focus on the sustainability of naturally-derived treatments that are often used by naturopaths, such herbal medicine [66,67], nutritional supplements and foods [68,69]. However, available literature assessing the intersection of climate change and sustainability of practice within the naturopathic profession has so far been limited to opinion pieces [41,70,71], clinical insight articles [42], conference proceedings [72], and localised research on naturopaths’ experiences of responding to climate change-related events [34]. The WHO has developed strategies for environmental sustainability in healthcare covering aspects such as resource and waste management, policy, workforce and community resilience [73], and communication about climate and health [40]. Considering that naturopathic organisations and committees have raised interest in the role of naturopathy regarding climate change and environmental sustainability [33,74,75], there may be untapped opportunities for naturopathic professionals to support development of a sustainable, resilient and climate change-ready profession that aligns with current global strategies for sustainability.

### Limitations

As always, the findings reported here should be viewed within the context of the study’s limitations. This is the first global study to date examining the perceptions and clinical behaviour of the naturopathic workforce with respect to climate change and environmental pollutants. Although some of the results can be correlated with other international studies, this study provides preliminary data and it is unknown whether they are entirely generalisable to the international naturopathic profession. The scope of this study and sub-group sample sizes did not permit inter-regional analysis; although a broad diversity of countries were represented, 40.2% of the respondents were from the Americas, hence the results may be skewed to this region. The recruitment frame being limited to members of professional associations and those naturopathic practitioners that follow the WNF social media may also bias the results toward those naturopaths who are more engaged in wider profession activity. Excepting the CCPS, the survey was not validated nor comprised of validated instruments, meaning care must be taken not to interpret the data beyond its descriptive intention. Some survey items required participants to report on patient characteristics and as the accuracy of this data cannot be independently confirmed, it is important this data is interpreted as solely representing practitioner perspectives of patient interactions (not the perspectives of patients themselves). Despite these limitations, this study offers an important foundational contribution of the role of the naturopathic workforce in addressing the health impacts of environmental pollutants and climate change.

## Conclusion

The naturopathic workforce demonstrates high awareness of climate change and environmental pollutants and their impacts on human health. While assessment and treatment practices frequently address food quality, access, and selected environmental exposures, other factors, particularly home, and workplace-based exposures and climate related stressors, are less consistently integrated into care. Naturopathic practitioners play a significant role in health literacy and education related to environmental health, and these findings highlight opportunities for enhanced training, resource development, and structural support to strengthen the integration of environmental and planetary health into clinical practice

## Declarations

### Ethics approval and consent to participate

Ethical approval was sought and obtained from the Human Research Ethics Committee at the University of Technology Sydney (ETH24-10002). Full study information and was provided to participants prior to recording consent and completing the survey.

### Consent for publication

No applicable

### Availability of data and materials

The dataset supporting the conclusions of this article is available in the Figshare repository, [embargoed pending publication: https://doi.org/10.6084/m9.figshare.32051841]

### Declaration of interest

The authors declare that they have no competing interests.

## Funding

No funding was received for this study.

### Authors’ contributions

HF, IL and MF contributed to the conception of the study, while HF, IL and AS developed the design and methodology. HF and IL contributed to the acquisition of the data, HF analysed the data, and all authors contributed to interpretation of the data. HF, IL and MF drafted the manuscript, and HF and AS substantively revised it. All authors have approved the final manuscript for submission.

## Supporting information

Supplemental File - Survey copy

## Data Availability

All data produced in the present study will be available upon publication at the Figshare online repository.

https://doi.org/10.6084/m9.figshare.32051841

## Acknowledgments

This survey was an initiative led by the WNF Environmental Health Committee which provided input into the scope of the survey. The authors are grateful to the individuals who generously volunteered their time to translate the survey and study materials. We would also like to thank WNF member organisations and the WNF social media community for distribution of the survey and support in recruitment for the survey.

## Author information

All authors have qualifications in naturopathy and roles with the WNF. During the conduct of this project, IL was the WNF Chief Executive Officer, MF and HF were Co-chairs of the Environmental Health Committee (now the Environmental Health Working Group), and AS was the Co-Chair of the Research Committee.

## List of abbreviations

CCPS: Climate Change Perception Scale
WHO: World Health Organization
WNF: World Naturopathic Federation

## Additional Information

### Additional File 1

A completed Strengthening the Reporting of Observational Studies in Epidemiology (STROBE) checklist is provided as Additional File 1.

File name: Additional File 1_STROBE checklist

File format: .pdf

Title of data: STROBE Checklist for manuscript ‘Climate change and environmental pollutants – an international survey of naturopathic perceptions and clinical behaviour’

### Additional File 2

The survey with participant information sheet is provided in all language translations in Additional File 2.

File name: Additional File 2_Full survey in 4 languages

File format: .pdf

Title of data: Survey instrument for study ‘Environmental Pollutants and Climate Change – an international survey of naturopathic perceptions and clinical behaviour’

Description of data: Participant information sheet and survey in all four languages.

## Notes

### Competing Interest Statement

The authors have declared no competing interest.

### Author Declarations

The Human Research Ethics Committee of the University of Technology Sydney gave ethical approval for this work (approval number: ETH24-10002).

