## Supplemental File - Survey copy for "Climate change and environmental pollutants – an international survey of naturopathic perceptions and clinical behaviour"

Additional File 2 – Survey instrument, Foley et. al.

Full survey in English 27 pages

Full survey in French 34 pages

Full survey in Portuguese 34 pages

Full survey in Spanish 35 pages

English

### Info sheet

#### Global Survey of the Naturopathic Profession on Climate Change and Environmental Health [ETH24-10002]

**WHO IS CONDUCTING THIS RESEARCH?**
This study is a collaboration between the World Naturopathic Federation (WNF) and Australian Research Consortium in Complementary and Integrative Medicine, University of Technology Sydney (UTS:ARCCIM).

My name is Dr Hope Foley and I am a researcher at UTS:ARCCIM and co-chair of the WNF Environmental Health Committee. I am conducting this research with Dr Iva Lloyd, CEO of the WNF, and research team members at UTS:ARCCIM, A/Prof Amie Steel, D/Prof Jon Adams, Dr Kirsten Baker and Mr Tristan Carter.

**WHAT IS THE RESEARCH ABOUT?**
The purpose of this research is to better understand the perspectives and practices of naturopathic practitioners around the world regarding the health impacts associated with environmental issues such as climate

change and environmental pollutants.

You have been invited to participate because you are a naturopathic practitioner who belongs to a professional association or organisation that is a member of the WNF (the association or organisation that contacted you about this research).

Before you decide to participate in this research study, please check the selection criteria. To participate in this study, you must:

- Have been active in clinical practice for at least 5 years
- Be able to complete a survey in one of the languages available (English, Spanish, Portuguese or French)

**FUNDING**

This project has not received any funding.

**WHAT DOES MY PARTICIPATION INVOLVE?**

Participation in this study is voluntary. It is completely up to you whether or not you decide to take part. If you decide to participate, we will invite you to complete an anonymous online survey that will take approximately 12 to 18 minutes to complete.

You can change your mind at any time and stop completing the survey/s without consequences. If you decide not to participate, or not to complete the survey, it will not affect your relationship with the researchers, UTS, the WNF, or the association/organisation that invited you. Participation is anonymous, so nobody will know if you participate or not.

**ARE THERE ANY RISKS/INCONVENIENCE?**

Yes, there are some risks/inconvenience. The time taken to complete the survey may be an inconvenience for some people. Some people may also experience the inconvenience of internet connectivity issues while trying to complete the questionnaire. In addition, the survey asks questions about climate change and environmental pollutants, which are topics that some people may find uncomfortable.

We have minimised these risks by keeping the survey as brief as possible and ensuring it functions on various device types. The questions that ask about climate change and environmental pollutants are not personal in nature, but related to your general perspectives and professional practices, to reduce any discomfort, and you will not be required to provide any information that you do not wish to share. If you experience discomfort or distress, please contact a suitable support person or healthcare professional.

**WHAT WILL HAPPEN TO INFORMATION ABOUT ME?**

The questionnaire is accessed via the button at the end of this information page. Completion of the survey is an indication of your consent. As the survey is anonymous, your data cannot be identified or removed after completion of the survey.

In accordance with relevant Australian and/or NSW Privacy laws, you have the right to request access to the information about you that is collected and stored by the research team. You also have the right to request that any information with which you disagree be corrected. Please inform the research team member named at the end of this document if you would like to access your information. This will only be possible if you provide personal information which may individually identify you, or be reasonably identifiable (e.g. if any open-text responses contextually identify or re-identify you).

It is anticipated that the results of this research project will be published and/or presented in a variety of forums, including WNF publications and presentations, and may be used in future projects conducted by the WNF. Data from the research will be stored in a secure location on UTS cloud-based platforms (e.g., OneDrive) accessible only to the research team. The results of this research may also be shared through open access (public) scientific databases, including internet databases. This will enable other researchers to use the data to investigate other important research questions. Results shared in this way will always be de-identified by removing all personal information (if you provide such information - e.g. your name, address, date of birth etc.) and/or any contextual information that could identify you.

**WHAT IF I HAVE ANY QUERIES OR CONCERNS?**
If you have any queries or concerns about the research that you think we can help you with, please feel free to contact me on. Alternatively, if you wish to speak with someone from the WNF, you may contact Dr Iva Lloyd on

If you would like to talk to someone who is not connected with the research, or if you have any concerns or complaints about any aspect of the conduct of this research that you wish to raise independently of the research team, please contact the Ethics Secretariat on +61 2 9514 2478 or and quote the UTS HREC reference number ETH24-10002. Any matter raised will be treated confidentially, investigated and you will be informed of the outcome.

**CONSENT**

Before you decide to participate in this research study, please double-check the selection criteria. To participate in this study, you must:

- Have been active in clinical practice for at least 5 years
- Be able to complete a survey in one of the languages available (English, Spanish, Portuguese or French)

○ Continue to survey
○ I do not wish to participate in the survey

#### Default Question Block

The following survey asks about your perspectives and experiences regarding aspects of climate change and environmental health in naturopathic practice. This survey will take approximately 15 minutes and your responses are completely anonymous.

#### Perceptions of climate and environmental health in the naturopathic community

The questions in this section ask about your broad perspectives on the topics of climate change and environmental health. We are interested in finding out what naturopathic clinicians think about these topics.

How much do you agree or disagree with the following statements?

| | Strongly agree | Agree | Neither agree nor disagree | Disagree | Strongly disagree |
|---|---|---|---|---|---|
| I believe that climate change is real | ○ | ○ | ○ | ○ | ○ |

| | Strongly agree | Agree | Neither agree nor disagree | Disagree | Strongly disagree |
|---|---|---|---|---|---|
| The main causes of climate change are human activities | ○ | ○ | ○ | ○ | ○ |
| Climate change will bring serious negative consequences | ○ | ○ | ○ | ○ | ○ |
| My local area will be influenced by climate change | ○ | ○ | ○ | ○ | ○ |
| It will be a long time before the consequences of climate change are felt | ○ | ○ | ○ | ○ | ○ |

| | Strongly agree | Agree | Neither agree nor disagree | Disagree | Strongly disagree |
|---|---|---|---|---|---|
| Climate change can affect human health | ○ | ○ | ○ | ○ | ○ |
| Climate change is already affecting human health | ○ | ○ | ○ | ○ | ○ |

#### How much do you agree or disagree with the following statements?

| | Strongly agree | Agree | Neither agree nor disagree | Disagree | Strongly disagree |
|---|---|---|---|---|---|
| I am concerned about the potential impacts of climate change on human health | ○ | ○ | ○ | ○ | ○ |
| I am concerned about the potential impacts of climate change on the health of my patients | ○ | ○ | ○ | ○ | ○ |

#### How much do you agree or disagree with the following statements?

| | Strongly agree | Agree | Neither agree nor disagree | Disagree | Strongly disagree |
|---|---|---|---|---|---|
| Environmental pollutants such as those found in the air, water, food or household environments are having negative impacts on human health | ○ | ○ | ○ | ○ | ○ |
| I am concerned about the effects of environmental pollutants on human health | ○ | ○ | ○ | ○ | ○ |
| I am concerned about the effects of environmental pollutants on the health of my patients | ○ | ○ | ○ | ○ | ○ |

#### How often do your patients express the following concerns about climate and environmental health?

| | Always | Most of the time | Sometimes | Rarely | Never |
|---|---|---|---|---|---|
| My patients express concerns about climate change impacting their physical health | ○ | ○ | ○ | ○ | ○ |

| | Always | Most of the time | Sometimes | Rarely | Never |
|---|---|---|---|---|---|
| My patients express concerns about climate change impacting their mental health | ○ | ○ | ○ | ○ | ○ |
| My patients express concerns about environmental pollutants or other environmental factors impacting their health | ○ | ○ | ○ | ○ | ○ |

#### Block 1

#### Climate and environmental health in naturopathic assessment

The following section includes questions about your experiences and perspectives regarding climate and environmental health in the context of naturopathic clinical assessment.

During the last 5 years, have you noticed any increase in how frequently you see the following symptoms and conditions in your clinical practice?

#### *Yes, I have noticed an increase in... (select all that apply)*

- [ ] Asthma, COPD and/or other respiratory conditions
- [ ] Cancer
- [ ] Dehydration conditions and/or heat stroke
- [ ] Kidney disease
- [ ] Anxiety, depression and/or other mental health concerns
- [ ] Neurological conditions
- [ ] Malnutrition concerns
- [ ] Food insecurity (low access to food)
- [ ] Mosquito-borne diseases
- [ ] Female infertility and reproductive dysfunction
- [ ] Male infertility and reproductive dysfunction
- [ ] Childhood development concerns
- [ ] Diabetes
- [ ] Autoimmune conditions
- [ ] None of the above

#### In your opinion, which of the following factors are contributing to this change/increase?

| | Climate change | Environmental pollutants | Economic factors | Social factors | Other | I don't know |
|---|---|---|---|---|---|---|
| » Asthma, COPD and/or other respiratory conditions | ☐ | ☐ | ☐ | ☐ | ☐ | ☐ |
| » Cancer | ☐ | ☐ | ☐ | ☐ | ☐ | ☐ |

| | Climate change | Environmental pollutants | Economic factors | Social factors | Other | I don't know |
|---|---|---|---|---|---|---|
| » Dehydration conditions and/or heat stroke | ☐ | ☐ | ☐ | ☐ | ☐ | ☐ |
| » Kidney disease | ☐ | ☐ | ☐ | ☐ | ☐ | ☐ |
| » Anxiety, depression and/or other mental health concerns | ☐ | ☐ | ☐ | ☐ | ☐ | ☐ |
| » Neurological conditions | ☐ | ☐ | ☐ | ☐ | ☐ | ☐ |
| » Malnutrition concerns | ☐ | ☐ | ☐ | ☐ | ☐ | ☐ |
| » Food insecurity (low access to food) | ☐ | ☐ | ☐ | ☐ | ☐ | ☐ |
| » Mosquito-borne diseases | ☐ | ☐ | ☐ | ☐ | ☐ | ☐ |
| » Female infertility and reproductive dysfunction | ☐ | ☐ | ☐ | ☐ | ☐ | ☐ |
| » Male infertility and reproductive dysfunction | ☐ | ☐ | ☐ | ☐ | ☐ | ☐ |
| » Childhood development concerns | ☐ | ☐ | ☐ | ☐ | ☐ | ☐ |
| » Diabetes | ☐ | ☐ | ☐ | ☐ | ☐ | ☐ |

| | Climate change | Environmental pollutants | Economic factors | Social factors | Other | I don't know |
|---|---|---|---|---|---|---|
| » Autoimmune conditions | ☐ | ☐ | ☐ | ☐ | ☐ | ☐ |
| » None of the above | ☐ | ☐ | ☐ | ☐ | ☐ | ☐ |

#### To what extent do you consider the following **climate-related factors** *during assessment of a patient's case*?

#### Food and water:

| | Always | Most of the time | Some of the time | Rarely | Never |
|---|---|---|---|---|---|
| The patient's **ability to access** fresh, healthy food | ○ | ○ | ○ | ○ | ○ |
| The environmental sustainability of patients' dietary habits (e.g., local sourcing, low carbon footprint | ○ | ○ | ○ | ○ | ○ |
| The patient's **ability to access** clean drinking water | ○ | ○ | ○ | ○ | ○ |
| Sufficient water intake for the patient's local climate and health status | ○ | ○ | ○ | ○ | ○ |

#### Air quality and weather:

| | Always | Most of the time | Some of the time | Rarely | Never |
|---|---|---|---|---|---|
| The patient's exposure to external air pollution from climate change or extreme weather-related events (e.g., smoke from forest fires) | ○ | ○ | ○ | ○ | ○ |
| The impact of weather changes or severe weather events on the patient's health and wellbeing | ○ | ○ | ○ | ○ | ○ |

To what extent do you consider the following **environmental factors** *during assessment of a patient's case?*

#### Food and water:

| | Always | Most of the time | Some of the time | Rarely | Never |
|---|---|---|---|---|---|
| **External** environmental factors affecting **food quality** for patients (e.g., organically grown foods, avoidance of pesticides or GMOs) | ○ | ○ | ○ | ○ | ○ |

| | Always | Most of the time | Some of the time | Rarely | Never |
|---|---|---|---|---|---|
| **Domestic** environmental factors affecting **food quality** for patients (e.g., toxins from food storage containers) | ○ | ○ | ○ | ○ | ○ |
| **External** environmental factors affecting patient's **water quality** (e.g., contamination of local water sources) | ○ | ○ | ○ | ○ | ○ |
| **Domestic** environmental factors affecting patient's **water quality** (e.g., filtration and storage) | ○ | ○ | ○ | ○ | ○ |

### Air quality and general environment:

| | Always | Most of the time | Some of the time | Rarely | Never |
|---|---|---|---|---|---|
| Patient's exposure to **external sources of air pollution** in living/working environments (e.g., car exhaust, industrial outputs) | ○ | ○ | ○ | ○ | ○ |

| | Always | Most of the time | Some of the time | Rarely | Never |
|---|---|---|---|---|---|
| Patient's exposure to **domestic/indoor sources of air pollution** in living/working environments (e.g., cleaning and personal care products, tobacco smoke, gas and wood-burning cooking appliances) | ○ | ○ | ○ | ○ | ○ |
| Patient's exposure to **other environmental toxins in the home, work or natural environments** (e.g., electromagnetic frequencies, light pollution) | ○ | ○ | ○ | ○ | ○ |
| **Testing patient** for presence of environmental toxic pollutants or associated impacts on health (e.g., testing for heavy metals and persistent organic pollutants) | ○ | ○ | ○ | ○ | ○ |

#### Block 2

### Naturopathic treatments and care for climate and environmental health

*The questions in this section ask about your experiences and perspectives on the types of care, prescriptions and treatments used by naturopaths in relation to climate and environmental health*

To what degree do you include the following options/factors when ***making recommendations or prescribing treatments*** to your patients?

| | Always | Most of the time | Some of the time | Rarely | Never |
|---|---|---|---|---|---|
| **Environmentally sustainable dietary and food-based** recommendations (e.g., locally sourced, seasonal, abundant supplies, low-carbon footprint) | ○ | ○ | ○ | ○ | ○ |
| **Environmentally sustainable herbs, supplements** and other prescribed remedies (e.g., locally sourced, seasonal, abundant supplies, low-carbon footprint) | ○ | ○ | ○ | ○ | ○ |
| The **quality of food** being recommended to or consumed by patients (e.g., organic, spray-free, farm-raised vs. wild-caught, plant-based) | ○ | ○ | ○ | ○ | ○ |

| | Always | Most of the time | Some of the time | Rarely | Never |
|---|---|---|---|---|---|
| Increasing **time that patients spend in nature** (nature prescribing) | ○ | ○ | ○ | ○ | ○ |
| Educating patients about the **connection between human health and the natural environment** | ○ | ○ | ○ | ○ | ○ |
| Educating patients about the **health impacts of environmental pollutants** | ○ | ○ | ○ | ○ | ○ |

When making recommendations or prescribing treatments to your patients, to what degree do you consider *decreasing the patient's exposure to environmental toxins in:*

| | Always | Most of the time | Some of the time | Rarely | Never |
|---|---|---|---|---|---|
| Food | ○ | ○ | ○ | ○ | ○ |
| Water | ○ | ○ | ○ | ○ | ○ |
| Air | ○ | ○ | ○ | ○ | ○ |
| Personal care products (e.g., bathing products, skincare, make-up) | ○ | ○ | ○ | ○ | ○ |

| | Always | Most of the time | Some of the time | Rarely | Never |
|---|---|---|---|---|---|
| Home-based products indoors (e.g., cleaning products, building materials) | ○ | ○ | ○ | ○ | ○ |
| Outdoor home-based products (e.g., gardening, pest control) | ○ | ○ | ○ | ○ | ○ |
| Workplace products, tools and environments | ○ | ○ | ○ | ○ | ○ |

#### Block 3

**Climate and environmental health in the broader naturopathic workforce**

The following questions ask about your thoughts on the broader role of naturopaths in the fields of climate change and environmental health.

Outside of clinical practice with individual patients, some practitioners engage in broader community education about health through **public platforms such as workshops, community talks, blogs/vlogs and social media**. To what extent do you engage in this type of public education about the following health topics?

| | Often | Sometimes | Occasionally | Rarely | Never |
|---|---|---|---|---|---|
| The connection between human health and the natural environment | ○ | ○ | ○ | ○ | ○ |
| The health impacts of climate change | ○ | ○ | ○ | ○ | ○ |
| The health impacts of environmental pollutants | ○ | ○ | ○ | ○ | ○ |
| Other health topics | ○ | ○ | ○ | ○ | ○ |

To what extent do you believe you, as a naturopath/ND, play a role in addressing the **health impacts of climate change** for your **individual patients**?

- ○ A very significant role
- ○ A significant role
- ○ A somewhat significant role
- ○ A slightly significant role
- ○ No role at all

To what extent do you believe you, as a naturopath/ND, play a role in addressing the **health impacts of climate change** for your **broader community**? (i.e., beyond the patients you see in clinic)

- ○ A very significant role

- A significant role
- A somewhat significant role
- A slightly significant role
- No role at all

How ***effective*** do you think naturopathic care is for addressing the **health impact of climate change**?

- Very effective
- Effective
- Somewhat effective
- Slightly effective
- Not effective at all

How ***confident*** do you feel about providing naturopathic care to address the **health impacts of climate change**?

- Very confident
- Confident
- Somewhat confident
- Not very confident
- Not confident at all

To what extent do you feel you need additional **education or training** on the topic of addressing the **health impacts of climate change**?

- Very strong need
- Strong need
- Some need
- Slight need
- I do not need any additional education or training

To what extent do you believe you, as a naturopath/ND, play a role in addressing the **health impacts of environmental toxins** for your **individual patients**?

- A very significant role
- A significant role
- A somewhat significant role
- A slightly significant role
- No role at all

To what extent do you believe you, as a naturopath/ND, play a role in addressing the **health impacts of environmental toxins** for your **broader community**? (i.e., beyond the patients you see in clinic)

- A very significant role

- A significant role
- A somewhat significant role
- A slightly significant role
- No role at all

How ***effective*** do you think naturopathic care is for addressing the **health impact of environmental toxins**?

- Very effective
- Effective
- Somewhat effective
- Slightly effective
- Not effective at all

How ***confident*** do you feel you feel about providing naturopathic care to address the **health impacts of environmental toxic pollutants**?

- Very confident
- Confident
- Somewhat confident
- Slightly confident
- Not confident at all

To what extent do you feel you need ***additional education or training*** on the topic of **environmental health and toxic pollutants**?

- Very strong need
- Strong need
- Some need
- Slight need
- I do not need any additional education or training

#### Block 4

#### About you

*To ensure we have included a diverse range of perspectives from naturopathic practitioners around the world, please provide a few details about yourself below. (Any information you provide here remains anonymous)*

In which country do you live?

Is this the same country in which your clinical practice is based?

- Yes
- No (please specify where you practice): ______

#### Which of the following best describes your gender?

- Female
- Male
- Non-binary
- Prefer to self-describe: ______

#### What was your age at your last birthday?

______

#### How many years have you been practicing as a naturopath/naturopathic doctor?

- Up to 10 years
- 11 to 15 years
- 16 to 20 years
- 21 to 25 years
- More than 25 years

Does your practice have a specialised focus? (select all that apply)

- ☐ No, my clinical practice has a ***general focus*** with no particular specialisation
- ☐ Yes, my clinical practice has a specialised focus on **environmental health**
- ☐ Yes, my clinical practice has a specialised focus on **climate-related health**
- ☐ Yes, my clinical practice has a specialised focus on **another topic** (please specify):

Which option best describes the clinical practice environment at your **primary place of practice**?

- ○ I am in a clinic by myself
- ○ I am in a clinic with other health professionals but no other naturopathic practitioners
- ○ I am in a clinic with other naturopathic practitioners but no other types of health professionals
- ○ I am in a clinic with other naturopathic practitioners and other health professionals
- ○ I am in a hospital setting
- ○ Other setting

Which of the below options best describes where your clinical practice is located?

☐ Urban or suburban area
☐ Regional area (neither urban nor rural)
☐ Rural or remote area
☐ Online

Powered by Qualtrics

Français

#### Info sheet

**Enquête mondiale sur le changement climatique et la santé environnementale auprès de la profession naturopathique [ETH24-10002]**

##### QUI MÈNE CETTE RECHERCHE ?

Cette étude est le fruit d'une collaboration entre la Fédération mondiale de naturopathie (WNF) et le Consortium Australien de Recherche en Médecine Complémentaire et Intégrative de l'Université de Technologie de Sydney (UTS:ARCCIM).

Je m'appelle Dr Hope Foley et je suis chercheuse à l'UTS : ARCCIM et coprésidente du Comité de santé environnementale de la WNF. Je mène cette recherche avec le Dr Iva Lloyd, PDG de la WNF, et les membres de l'équipe de recherche de l'UTS:ARCCIM, le professeur adjoint Amie Steel, le professeur adjoint Jon Adams, le Dr Kirsten Baker et M. Tristan Carter

##### SUR QUOI PORTE CETTE RECHERCHE ?

Le but de cette recherche est de mieux comprendre les perspectives et les pratiques des praticiens naturopathes du monde entier concernant les impacts sur la santé associés aux problèmes environnementaux tels que le changement climatique et les polluants environnementaux.

Vous avez été invité à participer parce que vous êtes un praticien naturopathes appartenant à une association ou organisation professionnelle membre de la WNF (l'association ou l'organisation qui vous a contacté au sujet de cette recherche).

**Avant de décider de participer à cette étude de recherche, merci de bien vérifier les critères de sélection. Pour participer à cette étude, vous devez** :

- Être actif en pratique clinique depuis au moins 5 ans
- Être capable de répondre à une enquête dans l'une des langues disponibles (anglais, espagnol, portugais ou français)

#### FINANCEMENT

Ce projet n'a reçu aucun financement.

#### EN QUOI CONSISTE MA PARTICIPATION ?

La participation à cette étude est volontaire. C'est à vous de décider si vous souhaitez y participer ou non. Si vous décidez de participer, nous vous inviterons à remplir une enquête en ligne anonyme qui prendra environ 12 à 18 minutes à compléter.

Vous pouvez changer d'avis à tout moment et arrêter de répondre à l'enquête/aux enquêtes sans conséquences. Si vous décidez de ne pas participer ou de ne pas terminer l'enquête, cela n'affectera pas votre relation avec les chercheurs, l'UTS, le WNF ou l'association/organisation qui vous a invité. La participation est anonyme, donc personne ne saura si vous participez ou non.

#### Y A-T-IL DES RISQUES/INCONVÉNIENTS ?

Oui, il existe certains risques/inconvénients. Le temps nécessaire pour répondre à l'enquête peut être un inconvénient pour certaines personnes. Certaines personnes peuvent également subir des problèmes de connectivité Internet lorsqu'elles tentent de remplir le questionnaire. En outre, l'enquête pose des questions sur le changement climatique et les polluants environnementaux, des sujets qui peuvent mettre certaines personnes mal à l'aise.

Nous avons minimisé ces risques en rendant l'enquête aussi brève que possible et en veillant à ce qu'elle fonctionne sur différents types d'appareils. Les questions sur le changement climatique et les polluants environnementaux ne sont pas de nature personnelle, mais liées à vos perspectives générales et à vos pratiques professionnelles, afin de réduire tout inconfort, vous ne serez pas tenu de fournir des informations que vous ne souhaitez pas partager. Si vous ressentez un inconfort ou une détresse, veuillez contacter une personne de soutien ou un professionnel de la santé approprié.

#### QU'ADVIENDRA-T-IL DES INFORMATIONS ME CONCERNANT ?

Le questionnaire est accessible via le bouton situé à la fin de cette page d'informations. Le fait de répondre à l'enquête constitue une indication de votre consentement. L'enquête étant anonyme, vos données ne peuvent pas être identifiées ou supprimées une fois l'enquête terminée.

Conformément aux lois australiennes et/ou de Nouvelle-Galles du Sud en vigueur en matière de confidentialité, vous avez le droit de demander l'accès aux informations vous concernant qui sont collectées et stockées par l'équipe de recherche. Vous avez également le droit de demander que toute information avec laquelle vous n'êtes pas d'accord soit corrigée. Veuillez informer le membre de l'équipe de recherche nommé à la fin de ce document si vous souhaitez accéder à vos informations. Cela ne sera possible que si vous fournissez des informations personnelles qui peuvent vous identifier individuellement ou être raisonnablement identifiables (par exemple, si des réponses en texte ouvert vous identifient ou vous réidentifient contextuellement).

Il est prévu que les résultats de ce projet de recherche soient publiés et/ou présentés dans divers forums, y compris les publications et présentations de la WNF, et pourront être utilisés dans de futurs projets menés par la WNF. Les données de la recherche seront stockées dans un emplacement sécurisé sur les plateformes cloud de l'UTS (par exemple, OneDrive) accessibles uniquement à l'équipe de recherche. Les résultats de cette recherche peuvent également être partagés via des bases de données scientifiques en libre accès (publiques), y compris des bases de données Internet. Cela permettra à d'autres chercheurs d'utiliser les données pour étudier d'autres questions de recherche importantes. Les résultats partagés de cette manière seront toujours anonymisés en supprimant toutes les informations personnelles (si vous fournissez de telles informations - par exemple votre nom, votre adresse, votre date de naissance, etc.) et/ou toute information contextuelle qui pourrait vous identifier

**QUE FAIRE SI J'AI DES QUESTIONS OU DES PRÉOCCUPATIONS ?**

Si vous avez des questions ou des préoccupations concernant la recherche pour lesquelles vous pensez que nous pouvons vous aider, n'hésitez pas à me contacter à l'adresse. Si vous souhaitez parler à quelqu'un de la WNF, vous pouvez contacter le Dr Iva Lloyd à l'adresse.

Si vous souhaitez parler à une personne qui n'est pas liée à la recherche, ou si vous avez des inquiétudes ou des plaintes concernant un aspect quelconque de la conduite de cette recherche que vous souhaitez les soulever indépendamment de l'équipe de recherche, veuillez contacter le Secrétariat d'éthique au +61 2 9514 2478 ou envoyer un e-mail à et indiquer le numéro de référence UTS HREC ETH24-10002. Toute question soulevée sera traitée de manière confidentielle, examinée et vous serez informé du résultat.

**CONSENTEMENT**

Avant de décider de participer à cette étude de recherche, veuillez vérifier les critères de sélection. Pour participer à cette étude, vous devez :

- Être actif en pratique clinique depuis au moins 5 ans
- Être capable de répondre à l’enquête dans l'une des langues disponibles (anglais, espagnol, portugais ou français)

### Enquête mondiale sur le changement climatique et la santé environnementale auprès de la profession naturopathique [ETH24-10002]

#### QUI MÈNE CETTE RECHERCHE ?

Cette étude est le fruit d'une collaboration entre la Fédération mondiale de naturopathie (WNF) et le Consortium Australien de Recherche en Médecine Complémentaire et Intégrative de l'Université de Technologie de Sydney (UTS:ARCCIM).

Je m'appelle Dr Hope Foley et je suis chercheuse à l'UTS : ARCCIM et coprésidente du Comité de santé environnementale de la WNF. Je mène cette recherche avec le Dr Iva Lloyd, PDG de la WNF, et les membres de l'équipe de recherche de l'UTS:ARCCIM, le professeur adjoint Amie Steel, le professeur adjoint Jon Adams, le Dr Kirsten Baker et M. Tristan Carter

#### SUR QUOI PORTE CETTE RECHERCHE ?

Le but de cette recherche est de mieux comprendre les perspectives et les pratiques des praticiens naturopathes du monde entier concernant les impacts sur la santé associés aux problèmes environnementaux tels que le changement climatique et les polluants environnementaux.

Vous avez été invité à participer parce que vous êtes un praticien naturopathes appartenant à une association ou organisation professionnelle membre de la WNF (l'association ou l'organisation qui vous a contacté au sujet de cette recherche).

**Avant de décider de participer à cette étude de recherche, merci de bien vérifier les critères de sélection. Pour participer à cette étude, vous devez** :

- Être actif en pratique clinique depuis au moins 5 ans
- Être capable de répondre à une enquête dans l'une des langues disponibles (anglais, espagnol, portugais ou français)

**FINANCEMENT**

Ce projet n'a reçu aucun financement.

**EN QUOI CONSISTE MA PARTICIPATION ?**

La participation à cette étude est volontaire. C'est à vous de décider si vous souhaitez y participer ou non. Si vous décidez de participer, nous vous inviterons à remplir une enquête en ligne anonyme qui prendra environ 12 à 18 minutes à compléter.

Vous pouvez changer d'avis à tout moment et arrêter de répondre à l'enquête/aux enquêtes sans conséquences. Si vous décidez de ne pas participer ou de ne pas terminer l'enquête, cela n'affectera pas votre relation avec les chercheurs, l'UTS, le WNF ou l'association/organisation qui vous a invité. La participation est anonyme, donc personne ne saura si vous participez ou non.

**Y A-T-IL DES RISQUES/INCONVÉNIENTS ?**

Oui, il existe certains risques/inconvénients. Le temps nécessaire pour répondre à l'enquête peut être un inconvénient pour certaines personnes. Certaines personnes peuvent également subir des problèmes de connectivité Internet lorsqu'elles tentent de remplir le questionnaire. En outre, l'enquête pose des questions sur le changement climatique et les polluants environnementaux, des sujets qui peuvent mettre certaines personnes mal à l'aise.

Nous avons minimisé ces risques en rendant l'enquête aussi brève que possible et en veillant à ce qu'elle fonctionne sur différents types d'appareils. Les questions sur le changement climatique et les polluants environnementaux ne sont pas de nature personnelle, mais liées à vos perspectives générales et à vos pratiques professionnelles, afin de réduire tout inconfort, vous ne serez pas tenu de fournir des informations que vous ne souhaitez pas partager. Si vous ressentez un inconfort ou une détresse, veuillez contacter une personne de soutien ou un professionnel de la santé approprié.

#### QU'ADVIENDRA-T-IL DES INFORMATIONS ME CONCERNANT ?

Le questionnaire est accessible via le bouton situé à la fin de cette page d'informations. Le fait de répondre à l'enquête constitue une indication de votre consentement. L'enquête étant anonyme, vos données ne peuvent pas être identifiées ou supprimées une fois l'enquête terminée.

Conformément aux lois australiennes et/ou de Nouvelle-Galles du Sud en vigueur en matière de confidentialité, vous avez le droit de demander l'accès aux informations vous concernant qui sont collectées et stockées par l'équipe de recherche. Vous avez également le droit de demander que toute information avec laquelle vous n'êtes pas d'accord soit corrigée. Veuillez informer le membre de l'équipe de recherche nommé à la fin de ce document si vous souhaitez accéder à vos informations. Cela ne sera possible que si vous fournissez des informations personnelles qui peuvent vous identifier individuellement ou être raisonnablement identifiables (par exemple, si des réponses en texte ouvert vous identifient ou vous réidentifient contextuellement).

Il est prévu que les résultats de ce projet de recherche soient publiés et/ou présentés dans divers forums, y compris les publications et présentations de la WNF, et pourront être utilisés dans de futurs projets menés par la WNF. Les données de la recherche seront stockées dans un emplacement sécurisé sur les plateformes cloud de l'UTS (par exemple, OneDrive) accessibles uniquement à l'équipe de recherche. Les résultats de cette recherche peuvent également être partagés via des bases de données scientifiques en libre accès (publiques), y compris des bases de données Internet. Cela permettra à d'autres chercheurs d'utiliser les données pour étudier d'autres questions de recherche importantes. Les résultats partagés de cette manière seront toujours anonymisés en supprimant toutes les informations personnelles (si vous fournissez de telles informations - par exemple votre nom, votre adresse, votre date de naissance, etc.) et/ou toute information contextuelle qui pourrait vous identifier

**QUE FAIRE SI J'AI DES QUESTIONS OU DES PRÉOCCUPATIONS ?**

Si vous avez des questions ou des préoccupations concernant la recherche pour lesquelles vous pensez que nous pouvons vous aider, n'hésitez pas à me contacter à l'adresse. Si vous souhaitez parler à quelqu'un de la WNF, vous pouvez contacter le Dr Iva

Lloyd à l'adresse.

Si vous souhaitez parler à une personne qui n'est pas liée à la recherche, ou si vous avez des inquiétudes ou des plaintes concernant un aspect quelconque de la conduite de cette recherche que vous souhaitez les soulever indépendamment de l'équipe de recherche, veuillez contacter le Secrétariat d'éthique au +61 2 9514 2478 ou envoyer un e-mail à et indiquer le numéro de référence UTS HREC ETH24-10002. Toute question soulevée sera traitée de manière confidentielle, examinée et vous serez informé du résultat.

**CONSENTEMENT**

Avant de décider de participer à cette étude de recherche, veuillez vérifier les critères de sélection. Pour participer à cette étude, vous devez :

- Être actif en pratique clinique depuis au moins 5 ans
- Être capable de répondre à l’enquête dans l'une des langues disponibles (anglais, espagnol, portugais ou français)

○ Participer à l'enquête

○ Je ne souhaite pas participer à l'enquête

#### Default Question Block

Le sondage suivant vous interroge sur vos points de vue et vos expériences concernant les aspects du changement climatique et de la santé environnementale dans la pratique naturopathique. Ce sondage prendra environ 15 minutes et vos réponses sont entièrement anonymes.

#### Perceptions du climat et de la santé environnementale dans la communauté naturopathique

Les questions de cette section portent sur vos perspectives générales sur les thèmes du changement climatique et de la santé environnementale. Nous souhaitons savoir ce que les cliniciens naturopathes pensent de ces sujets.

Dans quelle mesure êtes-vous d'accord ou en désaccord avec les affirmations suivantes ?

| | Tout à fait d'accord | D'accord | Neutre | Pas d'accord | Pas du tout d'accord |
|---|---|---|---|---|---|
| Je crois que le changement climatique est réel | ○ | ○ | ○ | ○ | ○ |

| | Tout à fait d'accord | D'accord | Neutre | Pas d'accord | Pas du tout d'accord |
|---|---|---|---|---|---|
| Les principales causes du changement climatique sont les activités humaines | ○ | ○ | ○ | ○ | ○ |
| Le changement climatique entraînera de graves conséquences négatives | ○ | ○ | ○ | ○ | ○ |
| Ma région sera influencée par le changement climatique | ○ | ○ | ○ | ○ | ○ |
| Il faudra beaucoup de temps avant que les conséquences du changement climatique ne se fassent sentir | ○ | ○ | ○ | ○ | ○ |
| Le changement climatique peut affecter la santé humaine | ○ | ○ | ○ | ○ | ○ |
| Le changement climatique affecte déjà la santé humaine | ○ | ○ | ○ | ○ | ○ |

Dans quelle mesure êtes-vous d’accord ou en désaccord avec les affirmations suivantes ?

| | Tout à fait d'accord | D'accord | Neutre | Pas d'accord | Pas du tout d'accord |
|---|---|---|---|---|---|
| Je suis préoccupé par les impacts potentiels du changement climatique sur la santé humaine | ○ | ○ | ○ | ○ | ○ |
| Je suis préoccupé par les impacts potentiels du changement climatique sur la santé de mes clients | ○ | ○ | ○ | ○ | ○ |

#### Dans quelle mesure êtes-vous d’accord ou en désaccord avec les affirmations suivantes ?

| | Tout à fait d'accord | D'accord | Neutre | Pas d'accord | Pas du tout d'accord |
|---|---|---|---|---|---|
| Les polluants environnementaux tels que ceux présents dans l’air, l’eau, les aliments ou les environnements domestiques ont des impacts négatifs sur la santé humaine | ○ | ○ | ○ | ○ | ○ |
| Je suis préoccupé par les effets des polluants environnementaux sur la santé humaine | ○ | ○ | ○ | ○ | ○ |

| | Tout à fait d'accord | D'accord | Neutre | Pas d'accord | Pas du tout d'accord |
|---|---|---|---|---|---|
| Je suis préoccupé par les effets des polluants environnementaux sur la santé de mes clients | ○ | ○ | ○ | ○ | ○ |

#### À quelle fréquence vos clients expriment-ils les préoccupations suivantes concernant le climat et la santé environnementale ?

| | Toujours | Souvent | Parfois | Rarement | Jamais |
|---|---|---|---|---|---|
| Mes clients expriment leurs inquiétudes quant à l’impact du changement climatique sur leur santé physique | ○ | ○ | ○ | ○ | ○ |
| Mes clients expriment leurs inquiétudes quant à l’impact du changement climatique sur leur santé mentale | ○ | ○ | ○ | ○ | ○ |

| | Toujours | Souvent | Parfois | Rarement | Jamais |
|---|---|---|---|---|---|
| Mes clients expriment des inquiétudes concernant les polluants environnementaux ou d’autres facteurs environnementaux ayant un impact sur leur santé | ◯ | ◯ | ◯ | ◯ | ◯ |

#### Block 1

##### Le climat et la santé environnementale dans l’évaluation naturopathique

La section suivante comprend des questions sur vos expériences et vos perspectives concernant le climat et la santé environnementale dans le contexte de l’évaluation clinique naturopathique.

Au cours des 5 dernières années, avez-vous remarqué une augmentation de la fréquence à laquelle vous observez les symptômes et affections suivants dans votre pratique clinique ?

#### *Oui, j'ai remarqué une augmentation de... (sélectionnez toutes les réponses applicables)*

☐ Asthme, BPCO et/ou autres affections respiratoires
☐ Cancer
☐ Conditions de déshydratation et/ou coup de chaleur
☐ Maladie du rein
☐ Anxiété, dépression et/ou autres problèmes de santé mentale
☐ Affections neurologiques
☐ Problèmes de malnutrition
☐ Insécurité alimentaire (faible accès à la nourriture)
☐ Maladies transmises par les moustiques
☐ Infertilité féminine et dysfonctionnement de la reproduction
☐ Infertilité masculine et dysfonctionnement de la reproduction
☐ Problèmes de développement de l’enfant
☐ Diabète
☐ Maladies auto-immunes
☐ Aucune des réponses ci-dessus

Selon vous, lesquels des facteurs suivants contribuent à ce changement/augmentation ?

| | Changement climatique | Polluants environnementaux | Facteurs économiques | Facteurs sociaux | Autre | [illegible] |
|---|---|---|---|---|---|---|
| » Asthme, BPCO et/ou autres affections respiratoires | ☐ | ☐ | ☐ | ☐ | ☐ | ☐ |

| | Changement climatique | Polluants environnementaux | Facteurs économiques | Facteurs sociaux | Autre | [illegible] |
|---|---|---|---|---|---|---|
| » Cancer | ☐ | ☐ | ☐ | ☐ | ☐ | ☐ |
| » Conditions de déshydratation et/ou coup de chaleur | ☐ | ☐ | ☐ | ☐ | ☐ | ☐ |
| » Maladie du rein | ☐ | ☐ | ☐ | ☐ | ☐ | ☐ |
| » Anxiété, dépression et/ou autres problèmes de santé mentale | ☐ | ☐ | ☐ | ☐ | ☐ | ☐ |
| » Affections neurologiques | ☐ | ☐ | ☐ | ☐ | ☐ | ☐ |
| » Problèmes de malnutrition | ☐ | ☐ | ☐ | ☐ | ☐ | ☐ |
| » Insécurité alimentaire (faible accès à la nourriture) | ☐ | ☐ | ☐ | ☐ | ☐ | ☐ |
| » Maladies transmises par les moustiques | ☐ | ☐ | ☐ | ☐ | ☐ | ☐ |
| » Infertilité féminine et dysfonctionnement de la reproduction | ☐ | ☐ | ☐ | ☐ | ☐ | ☐ |
| » Infertilité masculine et dysfonctionnement de la reproduction | ☐ | ☐ | ☐ | ☐ | ☐ | ☐ |

| | Changement climatique | Polluants environnementaux | Facteurs économiques | Facteurs sociaux | Autre | [illegible] |
|---|---|---|---|---|---|---|
| » Problèmes de développement de l'enfant | ☐ | ☐ | ☐ | ☐ | ☐ | ☐ |
| » Diabète | ☐ | ☐ | ☐ | ☐ | ☐ | ☐ |
| » Maladies auto-immunes | ☐ | ☐ | ☐ | ☐ | ☐ | ☐ |
| » Aucune des réponses ci-dessus | ☐ | ☐ | ☐ | ☐ | ☐ | ☐ |

Dans quelle mesure tenez-vous compte des **facteurs climatiques** suivants *lors de l'évaluation du cas d'un client* ?

#### Nourriture et eau :

| | Toujours | Souvent | Parfois | Rarement | Jamais |
|---|---|---|---|---|---|
| La **capacité** du client à accéder à des aliments frais et sains | ○ | ○ | ○ | ○ | ○ |

| | Toujours | Souvent | Parfois | Rarement | Jamais |
|---|---|---|---|---|---|
| La durabilité environnementale des habitudes alimentaires des clients (par exemple, approvisionnement local, faible empreinte carbone) | ○ | ○ | ○ | ○ | ○ |
| La **capacité** du client à accéder à l'eau potable | ○ | ○ | ○ | ○ | ○ |
| Consommation d'eau suffisante en fonction du climat local et de l'état de santé du client | ○ | ○ | ○ | ○ | ○ |

#### Qualité de l'air et météo :

| | Toujours | Souvent | Parfois | Rarement | Jamais |
|---|---|---|---|---|---|
| L'exposition du client à la pollution atmosphérique extérieure due au changement climatique ou à des phénomènes météorologiques extrêmes (par exemple, la fumée des incendies de forêt) | ○ | ○ | ○ | ○ | ○ |

| | Toujours | Souvent | Parfois | Rarement | Jamais |
|---|---|---|---|---|---|
| L'impact des changements climatiques ou des phénomènes météorologiques violents sur la santé et le bien-être du client | ○ | ○ | ○ | ○ | ○ |

#### Dans quelle mesure tenez-vous compte des **facteurs environnementaux** suivants *lors de l'évaluation du cas d'un client?*

#### Nourriture et eau :

| | Toujours | Souvent | Parfois | Rarement | Jamais |
|---|---|---|---|---|---|
| Facteurs environnementaux externes affectant la qualité des aliments pour les clients (par exemple, aliments cultivés de manière biologique, évitement des pesticides ou des OGM) | ○ | ○ | ○ | ○ | ○ |

| | Toujours | Souvent | Parfois | Rarement | Jamais |
|---|---|---|---|---|---|
| Facteurs environnementaux domestiques affectant la qualité des aliments pour les clients (p. ex. toxines provenant des contenants de stockage des aliments) | ○ | ○ | ○ | ○ | ○ |
| Facteurs environnementaux externes affectant la qualité de l'eau du client (par exemple, contamination des sources d'eau locales) | ○ | ○ | ○ | ○ | ○ |
| Facteurs environnementaux domestiques affectant la qualité de l'eau du client (par exemple, filtration et stockage) | ○ | ○ | ○ | ○ | ○ |

### Qualité de l'air et environnement général :

| | Toujours | Souvent | Parfois | Rarement | Jamais |
|---|---|---|---|---|---|
| Exposition du client à des **sources externes de pollution de l'air** dans les environnements de vie/travail (par exemple, gaz d'échappement des voitures, rejets industriels) | ○ | ○ | ○ | ○ | ○ |
| Exposition du client à des sources de pollution de l'air domestiques/intérieures dans les environnements de vie/de travail (par exemple, produits de nettoyage et de soins personnels, fumée de tabac, appareils de cuisson au gaz et au bois) | ○ | ○ | ○ | ○ | ○ |
| Exposition du client à d'autres toxines environnementales à la maison, au travail ou dans des environnements naturels (par exemple, fréquences électromagnétiques, pollution lumineuse) | ○ | ○ | ○ | ○ | ○ |
| **Test tester le patient** pour détecter la présence de polluants environnementaux toxiques ou d'impacts associés sur la santé (par exemple, dépistage de métaux lourds et de polluants organiques persistants) | ○ | ○ | ○ | ○ | ○ |

### Block 2

#### Traitements et soins naturopathiques pour la santé climatique et environnementale

*Les questions de cette section portent sur vos expériences et vos points de vue sur les types de soins, accompagnements proposés par les naturopathes en relation avec la santé climatique et environnementale*

Dans quelle mesure incluez-vous les options/facteurs suivants lorsque vous faites des recommandations à vos clients ?

| | Toujours | Souvent | Parfois | Rarement | Jamais |
|---|---|---|---|---|---|
| Recommandations **alimentaires et alimentaires respectueuses de l'environnement** (par exemple, d'origine locale, de saison, en abondance, à faible empreinte carbone) | ○ | ○ | ○ | ○ | ○ |

| | Toujours | Souvent | Parfois | Rarement | Jamais |
|---|---|---|---|---|---|
| **Herbes, suppléments et autres remèdes respectueux de l'environnement** (par exemple, d'origine locale, de saison, en abondance, à faible empreinte carbone) | ○ | ○ | ○ | ○ | ○ |
| La **qualité des aliments** recommandés aux clients ou consommés par eux (par exemple, biologiques, sans pulvérisation, élevés à la ferme ou pêchés dans la nature, à base de plantes) | ○ | ○ | ○ | ○ | ○ |
| Augmenter **le temps que les clients passent dans la nature** (prescription naturelle) | ○ | ○ | ○ | ○ | ○ |
| Éduquer les clients sur le **lien entre la santé humaine et l'environnement naturel** | ○ | ○ | ○ | ○ | ○ |
| Sensibiliser les clients aux **impacts des polluants environnementaux sur la santé** | ○ | ○ | ○ | ○ | ○ |

Lorsque vous faites des recommandations à vos clients, dans quelle mesure envisagez-vous de *diminuer l'exposition du client aux toxines environnementales dans :*

| | Toujours | Souvent | Parfois | Rarement | Jamais |
| --- | --- | --- | --- | --- | --- |
| Nourriture | ○ | ○ | ○ | ○ | ○ |
| Eau | ○ | ○ | ○ | ○ | ○ |
| Air | ○ | ○ | ○ | ○ | ○ |
| Produits de soins personnels (par exemple, produits cosmétiques, de bain, soins de la peau, maquillage) | ○ | ○ | ○ | ○ | ○ |
| Produits ménagers à l'intérieur (par exemple, produits de nettoyage, matériaux de construction) | ○ | ○ | ○ | ○ | ○ |
| Produits d'extérieur pour la maison (par exemple, jardinage, lutte antiparasitaire) | ○ | ○ | ○ | ○ | ○ |
| Produits, outils et environnements de travail | ○ | ○ | ○ | ○ | ○ |

### Block 3

**Santé climatique et environnementale dans le milieu naturopathique au sens large**

Les questions suivantes portent sur vos réflexions concernant le rôle plus large des naturopathes dans les domaines du changement climatique et de la santé environnementale.

En dehors de la pratique clinique avec des clients individuels, certains praticiens participent à une éducation communautaire plus large sur la santé par le biais de plateformes publiques telles que des ateliers, des discussions communautaires, des blogs/vlogs et des médias sociaux. Dans quelle mesure participez-vous à ce type d'éducation publique sur les sujets de santé suivants ?

| | Souvent | Parfois | Occasionnellement | Rarement | Jamais |
|---|---|---|---|---|---|
| Le lien entre la santé humaine et l'environnement naturel | ○ | ○ | ○ | ○ | ○ |
| Les impacts du changement climatique sur la santé | ○ | ○ | ○ | ○ | ○ |
| Les impacts des polluants environnementaux sur la santé | ○ | ○ | ○ | ○ | ○ |
| Autres sujets de santé | ○ | ○ | ○ | ○ | ○ |

Dans quelle mesure pensez-vous, en tant que naturopathe/naturopathe, jouer un rôle dans la lutte contre les **impacts du changement climatique** sur la santé de vos **clients** ?

- Un rôle très important
- Un rôle important
- Un rôle assez important
- Un rôle légèrement significatif
- Aucun rôle

Dans quelle mesure pensez-vous que vous, en tant que naturopathe/naturopathe, jouez un rôle dans la lutte contre les **impacts du changement climatique sur la santé** de votre **communauté au sens large** ? (c'est-à-dire au-delà des clients que vous voyez en clinique)

- Un rôle très important
- Un rôle important
- Un rôle assez important
- Un rôle légèrement significatif
- Aucun rôle

Dans quelle mesure pensez-vous que les soins naturopathiques sont efficaces pour lutter contre l'impact du changement climatique sur la santé ?

- Très efficace
- Efficace
- Plutôt efficace
- Légèrement efficace
- Pas efficace

Dans quelle mesure êtes-vous confiant quant à la possibilité de fournir des soins naturopathiques pour faire face aux **impacts du changement climatique sur la santé** ?

- Très confiant
- Confiant
- Plutôt confiant
- Pas très confiant
- Pas du tout confiant

Dans quelle mesure estimez-vous avoir besoin d'une **éducation ou d'une** formation supplémentaire sur le thème de la gestion des **impacts du changement climatique sur la santé** ?

- Un très fort besoin
- Un fort besoin
- Un certains besoin

- Un léger besoin
- Je n'ai pas besoin d'éducation ou de formation supplémentaire

Dans quelle mesure pensez-vous, en tant que naturopathe/médecin naturopathe, jouer un rôle dans la lutte contre les **impacts des toxines environnementales sur la santé** de vos **clients individuels** ?

- Un rôle très important
- Un rôle important
- Un rôle assez important
- Un rôle légèrement significatif
- Aucun rôle

Dans quelle mesure pensez-vous que vous, en tant que naturopathe/médecin naturopathe, jouez un rôle dans la lutte contre les **impacts des toxines environnementales sur la santé** de votre **communauté au sens large** ? (c'est-à-dire au-delà des clientsque vous voyez en clinique)

- Un rôle très important
- Un rôle important
- Un rôle assez important
- Un rôle légèrement significatif

- Aucun rôle

Dans quelle mesure pensez-vous que les soins naturopathiques sont efficaces pour lutter contre l'impact des toxines environnementales sur la santé ?

- Très efficace
- Efficace
- Plutôt efficace
- Légèrement efficace
- Pas efficace

Dans quelle mesure êtes-vous confiant quant à la possibilité de fournir des soins naturopathiques pour lutter contre les **impacts des polluants environnementaux toxiques** sur la santé ?

- Très confiant
- Confiant
- Plutôt confiant
- Légèrement confiant
- Pas du tout confiant

Dans quelle mesure estimez-vous avoir besoin d'une formation ou d'une éducation supplémentaire sur le thème de la **santé environnementale et des polluants toxiques** ?

- Un très fort besoin
- Un fort besoin
- Un certain besoin
- Un léger besoin
- Je n'ai pas besoin d'éducation ou de formation supplémentaire

#### Block 4

#### À propos de vous

*Pour nous assurer d'avoir inclus un large éventail de points de vue de praticiens naturopathes du monde entier, merci de nous foumir quelques détails sur vous-même ci-dessous. (Toutes les informations que vous fournissez ici restent anonymes)*

Dans quel pays vivez-vous ?

S'agit-il du même pays dans lequel votre pratique clinique est basée ?

- Oui

- ○ [ ] Non (veuillez préciser où vous exercez) :

#### Laquelle des propositions suivantes décrit le mieux votre genre ?

- ○ Femme
- ○ Homme
- ○ Non binaire
- ○ [ ] Je préfère me décrire moi-même :

#### Quel âge aviez-vous lors de votre dernier anniversaire ?

[ ]

#### Depuis combien d'années exercez-vous en tant que naturopathe/médecin naturopathe ?

- ○ Jusqu'à 10 ans
- ○ 11 à 15 ans
- ○ 16 à 20 ans
- ○ 21 à 25 ans
- ○ Plus de 25 ans

#### Votre cabinet est-il spécialisé ? (sélectionnez toutes les réponses appropriées)

- ☐ Non, ma pratique clinique est à orientation générale sans spécialisation particulière
- ☐ Oui, ma pratique clinique est spécialisée dans la **santé environnementale**
- ☐ Oui, ma pratique clinique est spécialisée dans la santé liée au climat.
- ☐ Oui, ma pratique clinique est spécialisée dans un **autre sujet** (veuillez préciser) :

#### Quelle option décrit le mieux l'environnement de pratique clinique de votre **lieu de pratique principal** ?

- ○ Je suis seul dans un cabinet
- ○ Je suis dans un cabinet avec d'autres professionnels de la santé mais pas d'autres praticiens en naturopathie
- ○ Je suis dans un cabinet avec d'autres praticiens naturopathes mais aucun autre type de professionnels de la santé
- ○ Je suis dans un cabinet avec d'autres praticiens naturopathes et d'autres professionnels de la santé
- ○ Je suis dans un milieu hospitalier
- ○ Autre paramètre

Laquelle des options ci-dessous décrit le mieux l’endroit où se situe votre pratique clinique ?

- ☐ Zone urbaine ou péri-urbaine
- ☐ Zone régionale (ni urbaine, ni rurale)
- ☐ Zone rurale ou éloignée
- ☐ En ligne

Powered by Qualtrics

Português

#### Info sheet

**Pesquisa global da profissão naturopática sobre mudanças climáticas e saúde ambiental[ETH24-10002]**

**QUEM ESTÁ CONDUZINDO ESTA PESQUISA?**
Este estudo é uma colaboração entre a World Naturopathic Federation (WNF) e o Australian Research Consortium in Complementary and Integrative Medicine, University of Technology Sydney (UTS:ARCCIM).

Meu nome é Dra. Hope Foley e sou pesquisadora na UTS:ARCCIM e copresidente do Comitê de Saúde Ambiental da WNF. Estou conduzindo esta pesquisa com a Dra. Iva Lloyd, CEO da WNF, e membros da equipe de pesquisa da UTS:ARCCIM, Profa. Assoc. Amie Steel, Prof. Distinto Jon Adams, Dra. Kirsten Baker e Sr. Tristan Carter.

**SOBRE O QUE É A PESQUISA?**
O objetivo desta pesquisa é entender melhor as perspectivas e práticas de praticantes naturopatas ao redor do mundo em relação aos impactos na saúde associados a questões ambientais, como mudanças climáticas e poluentes ambientais.

Você foi convidado a participar porque é um praticante naturopata que pertence a uma associação ou organização profissional que é membro da WNF (a associação ou organização que o contatou sobre esta pesquisa).

Antes de decidir participar deste estudo de pesquisa, verifique os critérios de seleção. Para participar deste estudo, você deve:

- Ter atuado na prática clínica por pelo menos 5 anos
- Ser capaz de concluir uma pesquisa em um dos idiomas disponíveis (inglês, espanhol, português ou francês)

**FINANCIAMENTO**

Este projeto não recebeu nenhum financiamento.

**O QUE ENVOLVE MINHA PARTICIPAÇÃO?**

A participação neste estudo é voluntária. Depende totalmente de você decidir participar ou não. Se você decidir participar, convidaremos você a completar uma pesquisa anônima on-line que levará aproximadamente de 12 a 18 minutos para ser concluída.

Você pode mudar de ideia a qualquer momento e parar de completar a(s) pesquisa(s) sem consequências. Se você decidir não participar ou não completar a pesquisa, isso não afetará seu relacionamento com os pesquisadores, a UTS, a WNF ou a associação/organização que o convidou. A participação é anônima, então ninguém saberá se você participa ou não.

**EXISTEM RISCOS/INCONVENIÊNCIAS?**

Sim, existem alguns riscos/inconveniências. O tempo gasto para completar a pesquisa pode ser um inconveniente para algumas pessoas. Algumas pessoas também podem experimentar o inconveniente de problemas de conectividade com a Internet ao tentar completar o questionário. Além disso, a pesquisa faz perguntas sobre mudanças climáticas e poluentes ambientais, que são tópicos que algumas pessoas podem achar desconfortáveis.

Minimizamos esses riscos mantendo a pesquisa o mais breve possível e garantindo que ela funcione em vários tipos de dispositivos. As perguntas sobre mudanças climáticas e poluentes ambientais não são de natureza pessoal, mas relacionadas às suas perspectivas gerais e práticas profissionais, para reduzir qualquer desconforto, e você não será obrigado a fornecer nenhuma informação que não deseja compartilhar. Se sentir desconforto ou angústia, entre em contato com uma pessoa de suporte ou profissional de saúde adequado.

#### O QUE ACONTECERÁ COM AS INFORMAÇÕES SOBRE MIM?

O questionário é acessado por meio do botão no final desta página de informações. A conclusão da pesquisa é uma indicação do seu consentimento. Como a pesquisa é anônima, seus dados não podem ser identificados ou removidos após a conclusão da pesquisa.

De acordo com as leis de privacidade relevantes da Austrália e/ou NSW, você tem o direito de solicitar acesso às informações sobre você que são coletadas e armazenadas pela equipe de pesquisa. Você também tem o direito de solicitar que qualquer informação com a qual você discorde seja corrigida. Informe o membro da equipe de pesquisa nomeado no final deste documento se você gostaria de acessar suas informações. Isso só será possível se você fornecer informações pessoais que possam identificá-lo individualmente ou ser razoavelmente identificáveis (por exemplo, se alguma resposta de texto aberto o identificar ou reidentificar contextualmente).

É previsto que os resultados deste projeto de pesquisa sejam publicados e/ou apresentados em uma variedade de fóruns, incluindo publicações e apresentações da WNF, e podem ser usados em projetos futuros conduzidos pela WNF. Os resultados desta pesquisa também podem ser compartilhados por meio de bancos de dados científicos de acesso aberto (públicos), incluindo bancos de dados da Internet. Isso permitirá que outros pesquisadores usem os dados para investigar outras questões importantes de pesquisa. Os resultados compartilhados dessa forma sempre serão desidentificados, removendo todas as informações pessoais (se você fornecer tais informações - por exemplo, seu nome, endereço, data de nascimento etc.) e/ou quaisquer informações contextuais que possam identificá-lo.

**E SE EU TIVER ALGUMA DÚVIDA OU PREOCUPAÇÃO?**
Se você tiver alguma dúvida ou preocupação sobre a pesquisa com a qual acha que podemos ajudá-lo, sinta-se à vontade para entrar em contato comigo em. Alternativamente, se você deseja falar com alguém da WNF, você pode contatar a Dra. Iva Lloyd em

Se você gostaria de falar com alguém que não esteja conectado com a pesquisa, ou se você tiver alguma preocupação ou reclamação sobre qualquer aspecto da condução desta pesquisa que você deseja levantar independentemente da equipe de pesquisa, por favor, entre em contato com a Secretaria de Ética em +61 2 9514 2478 ou envie um e-mail para e cite o número de referência UTS HREC ETH24-10002. Qualquer questão levantada será tratada confidencialmente, investigada e você será informado do resultado.

**CONSENTIMENTO**
Antes de decidir participar deste estudo de pesquisa, por favor, verifique novamente os critérios de seleção. Para participar deste estudo, você deve:

- Ter atuado na prática clínica por pelo menos 5 anos
- Ser capaz de responder a uma pesquisa em um dos idiomas disponíveis (inglês, espanhol, português, francês ou italiano)

#### Pesquisa global da profissão naturopática sobre mudanças climáticas e saúde ambiental[ETH24-10002]

**QUEM ESTÁ CONDUZINDO ESTA PESQUISA?**
Este estudo é uma colaboração entre a World Naturopathic Federation (WNF) e o Australian Research Consortium in Complementary and Integrative Medicine, University of Technology Sydney (UTS:ARCCIM).

Meu nome é Dra. Hope Foley e sou pesquisadora na UTS:ARCCIM e copresidente do Comitê de Saúde Ambiental da WNF. Estou conduzindo esta pesquisa com a Dra. Iva Lloyd, CEO da WNF, e membros da equipe de pesquisa da UTS:ARCCIM, Profa. Assoc. Amie Steel, Prof. Distinto Jon Adams, Dra. Kirsten Baker e Sr. Tristan Carter.

**SOBRE O QUE É A PESQUISA?**

O objetivo desta pesquisa é entender melhor as perspectivas e práticas de praticantes naturopatas ao redor do mundo em relação aos impactos na saúde associados a questões ambientais, como mudanças climáticas e poluentes ambientais.

Você foi convidado a participar porque é um praticante naturopata que pertence a uma associação ou organização profissional que é membro da WNF (a associação ou organização que o contatou sobre esta pesquisa).

Antes de decidir participar deste estudo de pesquisa, verifique os critérios de seleção. Para participar deste estudo, você deve:

- Ter atuado na prática clínica por pelo menos 5 anos
- Ser capaz de concluir uma pesquisa em um dos idiomas disponíveis (inglês, espanhol, português ou francês)

**FINANCIAMENTO**

Este projeto não recebeu nenhum financiamento.

**O QUE ENVOLVE MINHA PARTICIPAÇÃO?**

A participação neste estudo é voluntária. Depende totalmente de você decidir participar ou não. Se você decidir participar, convidaremos você a completar uma pesquisa anônima on-line que levará aproximadamente de 12 a 18 minutos para ser concluída.

Você pode mudar de ideia a qualquer momento e parar de completar a(s) pesquisa(s) sem consequências. Se você decidir não participar ou não completar a pesquisa, isso não afetará seu relacionamento com os pesquisadores, a UTS, a WNF ou a associação/organização que o convidou. A participação é anônima, então ninguém saberá se você participa ou não.

**EXISTEM RISCOS/INCONVENIÊNCIAS?**

Sim, existem alguns riscos/inconveniências. O tempo gasto para completar a pesquisa pode ser um inconveniente para algumas pessoas. Algumas pessoas também podem experimentar o inconveniente de problemas de conectividade com a Internet ao tentar completar o questionário. Além disso, a pesquisa faz perguntas sobre mudanças climáticas e poluentes ambientais, que são tópicos que algumas pessoas podem achar desconfortáveis.

Minimizamos esses riscos mantendo a pesquisa o mais breve possível e garantindo que ela funcione em vários tipos de dispositivos. As perguntas sobre mudanças climáticas e poluentes ambientais não são de natureza pessoal, mas relacionadas às suas perspectivas gerais e práticas profissionais, para reduzir qualquer desconforto, e você não será obrigado a fornecer nenhuma informação que não deseja compartilhar. Se sentir desconforto ou angústia, entre em contato com uma pessoa de suporte ou profissional de saúde adequado.

#### O QUE ACONTECERÁ COM AS INFORMAÇÕES SOBRE MIM?

O questionário é acessado por meio do botão no final desta página de informações. A conclusão da pesquisa é uma indicação do seu consentimento. Como a pesquisa é anônima, seus dados não podem ser identificados ou removidos após a conclusão da pesquisa.

De acordo com as leis de privacidade relevantes da Austrália e/ou NSW, você tem o direito de solicitar acesso às informações sobre você que são coletadas e armazenadas pela equipe de pesquisa. Você também tem o direito de solicitar que qualquer informação com a qual você discorde seja corrigida. Informe o membro da equipe de pesquisa nomeado no final deste documento se você gostaria de acessar suas informações. Isso só será possível se você fornecer informações pessoais que possam identificá-lo individualmente ou ser razoavelmente identificáveis (por exemplo, se alguma resposta de texto aberto o identificar ou reidentificar contextualmente).

É previsto que os resultados deste projeto de pesquisa sejam publicados e/ou apresentados em uma variedade de fóruns, incluindo publicações e apresentações da WNF, e podem ser usados em projetos futuros conduzidos pela WNF. Os resultados desta pesquisa também podem ser compartilhados por meio de bancos de dados científicos de acesso aberto (públicos), incluindo bancos de dados da Internet. Isso permitirá que outros pesquisadores usem os dados para investigar outras questões importantes de pesquisa. Os resultados compartilhados dessa forma sempre serão desidentificados, removendo todas as informações pessoais (se você fornecer tais informações - por exemplo, seu nome, endereço, data de nascimento etc.) e/ou quaisquer informações contextuais que possam identificá-lo.

**E SE EU TIVER ALGUMA DÚVIDA OU PREOCUPAÇÃO?**
Se você tiver alguma dúvida ou preocupação sobre a pesquisa com a qual acha que podemos ajudá-lo, sinta-se à vontade para entrar em contato comigo em. Alternativamente, se você deseja falar com alguém da WNF, você pode contatar a Dra. Iva Lloyd em

Se você gostaria de falar com alguém que não esteja conectado com a pesquisa, ou se você tiver alguma preocupação ou reclamação sobre qualquer aspecto da condução desta pesquisa que você deseja levantar independentemente da equipe de pesquisa, por favor, entre em contato com a Secretaria de Ética em +61 2 9514 2478 ou envie um e-mail para e cite o número de referência UTS HREC ETH24-10002. Qualquer questão levantada será tratada confidencialmente, investigada e você será informado do resultado.

**CONSENTIMENTO**
Antes de decidir participar deste estudo de pesquisa, por favor, verifique novamente os critérios de seleção. Para participar deste estudo, você deve:

- Ter atuado na prática clínica por pelo menos 5 anos
- Ser capaz de responder a uma pesquisa em um dos idiomas disponíveis (inglês, espanhol, português, francês ou italiano)

○ Continuar a pesquisa
○ Não desejo participar da pesquisa

#### Default Question Block

A pesquisa a seguir pergunta sobre suas perspectivas e experiências em relação a aspectos de mudança climática e saúde ambiental na prática naturopática. Esta pesquisa levará aproximadamente 15 minutos e suas respostas são completamente anônimas.

#### Percepções sobre o clima e a saúde ambiental na comunidade naturopática

As perguntas nesta seção perguntam sobre suas perspectivas gerais sobre os tópicos de mudança climática e saúde ambiental. Estamos interessados em descobrir o que os clínicos naturopatas pensam sobre esses tópicos.

O quanto você concorda ou discorda das seguintes afirmações?

| | Concordo totalmente | Concordo | Nem concordo nem discordo | Discordo | Discordo totalmente |
|---|---|---|---|---|---|
| Acredito que a mudança climática é real | ○ | ○ | ○ | ○ | ○ |

| | Concordo totalmente | Concordo | Nem concordo nem discordo | Discordo | Discordo totalmente |
|---|---|---|---|---|---|
| As principais causas das mudanças climáticas são as atividades humanas | ○ | ○ | ○ | ○ | ○ |
| As mudanças climáticas trarão consequências negativas graves | ○ | ○ | ○ | ○ | ○ |
| Minha região será influenciada pelas mudanças climáticas | ○ | ○ | ○ | ○ | ○ |
| Vai demorar muito tempo até que as consequências das mudanças climáticas sejam sentidas | ○ | ○ | ○ | ○ | ○ |
| As mudanças climáticas podem afetar a saúde humana | ○ | ○ | ○ | ○ | ○ |
| As mudanças climáticas já estão a afetando a saúde humana | ○ | ○ | ○ | ○ | ○ |

#### O quanto você concorda ou discorda das seguintes afirmações?

| | Concordo totalmente | Concordo | Nem concordo nem discordo | Discordo | Discordo totalmente |
|---|---|---|---|---|---|
| Estou preocupado com os potenciais impactos das mudanças climáticas na saúde humana | ○ | ○ | ○ | ○ | ○ |
| Estou preocupado com os potenciais impactos das mudanças climáticas na saúde dos meus pacientes | ○ | ○ | ○ | ○ | ○ |

#### O quanto você concorda ou discorda das seguintes afirmações?

| | Concordo totalmente | Concordo | Nem concordo nem discordo | Discordo | Discordo totalmente |
|---|---|---|---|---|---|
| Poluentes ambientais, como os encontrados no ar, na água, nos alimentos ou nos ambientes domésticos, estão tendo impactos negativos na saúde humana | ○ | ○ | ○ | ○ | ○ |
| Estou preocupado com os efeitos dos poluentes ambientais na saúde humana | ○ | ○ | ○ | ○ | ○ |

| | Concordo totalmente | Concordo | Nem concordo nem discordo | Discordo | Discordo totalmente |
|---|---|---|---|---|---|
| Estou preocupado com os efeitos dos poluentes ambientais na saúde dos meus pacientes | ○ | ○ | ○ | ○ | ○ |

#### Com que frequência seus pacientes expressam as seguintes preocupações sobre o clima e a saúde ambiental?

| | Sempre | Na maioria das vezes | Às vezes | Raramente | Nunca |
|---|---|---|---|---|---|
| Meus pacientes expressam preocupações sobre o impacto das mudanças climáticas em sua saúde física | ○ | ○ | ○ | ○ | ○ |
| Meus pacientes expressam preocupações sobre o impacto das mudanças climáticas em sua saúde mental | ○ | ○ | ○ | ○ | ○ |

| | Sempre | Na maioria das vezes | Às vezes | Raramente | Nunca |
|---|---|---|---|---|---|
| Meus pacientes expressam preocupações sobre poluentes ambientais ou outros fatores ambientais que afetam sua saúde | ○ | ○ | ○ | ○ | ○ |

#### Block 1

#### Saúde climática e ambiental na avaliação naturopática

A seção a seguir inclui perguntas sobre suas experiências e perspectivas em relação à saúde climática e ambiental no contexto da avaliação clínica naturopática.

Durante os últimos 5 anos, você notou algum aumento na frequência com que vê os seguintes sintomas e condições em sua prática clínica?

***Sim, notei um aumento em... (selecione todas as opções aplicáveis)***

- ☐ Asma, DPOC e/ou outras doenças respiratórias
- ☐ Câncer
- ☐ Condições de desidratação e/ou insolação
- ☐ Doença renal
- ☐ Ansiedade, depressão e/ou outros problemas de saúde mental
- ☐ Condições neurológicas
- ☐ Sinais de desnutrição
- ☐ Insegurança alimentar (baixo acesso a alimentos)
- ☐ Doenças transmitidas por mosquitos
- ☐ Infertilidade feminina e disfunção reprodutiva
- ☐ Infertilidade masculina e disfunção reprodutiva
- ☐ Questões relacionadas ao desenvolvimento infantil
- ☐ Diabetes
- ☐ Condições autoimunes
- ☐ Nenhuma das acima

Na sua opinião, quais dos seguintes fatores estão contribuindo para essa mudança/aumento?

| | Mudanças climáticas | Poluentes ambientais | Fatores econômicos | Fatores sociais | Outro | Não sei |
|---|---|---|---|---|---|---|
| » Asma, DPOC e/ou outras doenças respiratórias | ☐ | ☐ | ☐ | ☐ | ☐ | ☐ |
| » Câncer | ☐ | ☐ | ☐ | ☐ | ☐ | ☐ |
| » Condições de desidratação e/ou insolação | ☐ | ☐ | ☐ | ☐ | ☐ | ☐ |

| | Mudanças climáticas | Poluentes ambientais | Fatores econômicos | Fatores sociais | Outro | Não sei |
|---|---|---|---|---|---|---|
| » Doença renal | ☐ | ☐ | ☐ | ☐ | ☐ | ☐ |
| » Ansiedade, depressão e/ou outros problemas de saúde mental | ☐ | ☐ | ☐ | ☐ | ☐ | ☐ |
| » Condições neurológicas | ☐ | ☐ | ☐ | ☐ | ☐ | ☐ |
| » Sinais de desnutrição | ☐ | ☐ | ☐ | ☐ | ☐ | ☐ |
| » Insegurança alimentar (baixo acesso a alimentos) | ☐ | ☐ | ☐ | ☐ | ☐ | ☐ |
| » Doenças transmitidas por mosquitos | ☐ | ☐ | ☐ | ☐ | ☐ | ☐ |
| » Infertilidade feminina e disfunção reprodutiva | ☐ | ☐ | ☐ | ☐ | ☐ | ☐ |
| » Infertilidade masculina e disfunção reprodutiva | ☐ | ☐ | ☐ | ☐ | ☐ | ☐ |
| » Questões relacionadas ao desenvolvimento infantil | ☐ | ☐ | ☐ | ☐ | ☐ | ☐ |
| » Diabetes | ☐ | ☐ | ☐ | ☐ | ☐ | ☐ |

| | Mudanças climáticas | Poluentes ambientais | Fatores econômicos | Fatores sociais | Outro | Não sei |
|---|---|---|---|---|---|---|
| » Condições autoimunes | ☐ | ☐ | ☐ | ☐ | ☐ | ☐ |
| » Nenhuma das acima | ☐ | ☐ | ☐ | ☐ | ☐ | ☐ |

Até que ponto você considera os seguintes **fatores relacionados ao clima** *durante a avaliação do caso de um paciente*?

#### Comida e água:

| | Sempre | Na maioria das vezes | Algumas vezes | Raramente | Nunca |
|---|---|---|---|---|---|
| A **capacidade** do paciente de ter acesso a alimentos frescos e saudáveis | ○ | ○ | ○ | ○ | ○ |
| A sustentabilidade ambiental dos hábitos alimentares dos pacientes (por exemplo, abastecimento local, baixa pegada de carbono | ○ | ○ | ○ | ○ | ○ |
| A **capacidade** do paciente de ter acesso a água potável limpa | ○ | ○ | ○ | ○ | ○ |

| | Sempre | Na maioria das vezes | Algumas vezes | Raramente | Nunca |
|---|---|---|---|---|---|
| Ingestão de água suficiente para o clima local e estado de saúde do paciente | ○ | ○ | ○ | ○ | ○ |

#### Qualidade do ar e clima:

| | Sempre | Na maioria das vezes | Algumas vezes | Raramente | Nunca |
|---|---|---|---|---|---|
| A exposição do paciente à poluição atmosférica externa causada por mudanças climáticas ou eventos climáticos extremos (por exemplo, fumaca de incêndios florestais) | ○ | ○ | ○ | ○ | ○ |
| O impacto das mudanças climáticas ou eventos climáticos severos na saúde e bem-estar do paciente | ○ | ○ | ○ | ○ | ○ |

### Até que ponto você considera os seguintes **fatores ambientais** *durante a avaliação do caso de um paciente?*

### Comida e água:

| | Sempre | Na maioria das vezes | Algumas vezes | Raramente | Nunca |
|---|---|---|---|---|---|
| **Fatores ambientais externos que afetam a qualidade** dos alimentos para os pacientes (por exemplo, alimentos cultivados organicamente, evitar pesticidas ou alimentos geneticamente modificados) | ○ | ○ | ○ | ○ | ○ |
| **Fatores ambientais** domésticos que afetam a **qualidade** dos alimentos para os pacientes (por exemplo, toxinas de recipientes de armazenamento de alimentos) | ○ | ○ | ○ | ○ | ○ |
| **Fatores ambientais externos que afetam a qualidade da água** do paciente (por exemplo, contaminação de fontes de água locais) | ○ | ○ | ○ | ○ | ○ |

| | Sempre | Na maioria das vezes | Algumas vezes | Raramente | Nunca |
|---|---|---|---|---|---|
| **Fatores ambientais** domésticos que afetam a **qualidade da água** do paciente (por exemplo, filtragem e armazenamento) | ○ | ○ | ○ | ○ | ○ |

#### Qualidade do ar e ambiente geral:

| | Sempre | Na maioria das vezes | Algumas vezes | Raramente | Nunca |
|---|---|---|---|---|---|
| Exposição do paciente a **fontes externas de poluição do ar** em ambientes de vida/trabalho (por exemplo, escapamento de automóveis, emissões industriais) | ○ | ○ | ○ | ○ | ○ |
| Exposição do paciente a fontes domésticas/internas de poluição do ar em ambientes de vida/trabalho (por exemplo, produtos de limpeza e cuidados pessoais, fumaça de tabaco, aparelhos de cozinha a gás e a lenha) | ○ | ○ | ○ | ○ | ○ |

| | Sempre | Na maioria das vezes | Algumas vezes | Raramente | Nunca |
|---|---|---|---|---|---|
| Exposição do paciente a **outras toxinas ambientais em casa, no trabalho ou em ambientes naturais** (por exemplo, frequências eletromagnéticas, poluição luminosa) | ○ | ○ | ○ | ○ | ○ |
| **Teste do paciente** para presença de poluentes tóxicos ambientais ou impactos associados à saúde (por exemplo, teste de metais pesados e poluentes orgânicos persistentes) | ○ | ○ | ○ | ○ | ○ |

#### Block 2

### Tratamentos naturopáticos e cuidados para a saúde climática e ambiental

*As perguntas nesta seção perguntam sobre suas experiências e perspectivas sobre os tipos de cuidados, prescrições e tratamentos usados por naturopatas em relação à saúde climática e ambiental*

Até que ponto você inclui as seguintes opções/fatores ao ***fazer recomendações ou prescrever tratamentos*** aos seus pacientes?

| | Sempre | Na maioria das vezes | Algumas vezes | Raramente | Nunca |
|---|---|---|---|---|---|
| **Recomendações alimentares e alimento ambientalmente sustentáveis** (por exemplo, de origem local, sazonais, de oferta abundante, baixa pegada de carbono) | ○ | ○ | ○ | ○ | ○ |
| **Ervas, suplementos e outros remédios prescritos ambientalmente sustentáveis** (por exemplo, de origem local, sazonais, de oferta abundante, baixa pegada de carbono) | ○ | ○ | ○ | ○ | ○ |
| A **qualidade** dos alimentos recomendados ou consumidos pelos pacientes (por exemplo, orgânicos, sem pulverização, criados em fazendas ou selvagens, à base de plantas) | ○ | ○ | ○ | ○ | ○ |

| | Sempre | Na maioria das vezes | Algumas vezes | Raramente | Nunca |
|---|---|---|---|---|---|
| Aumentar o tempo que os pacientes passam na natureza (prescrição natureza) | ○ | ○ | ○ | ○ | ○ |
| Educar os pacientes sobre a **conexão entre a saúde humana e o ambiente natural** | ○ | ○ | ○ | ○ | ○ |
| Educar os pacientes sobre os **impactos dos poluentes ambientais na saúde** | ○ | ○ | ○ | ○ | ○ |

#### Ao fazer recomendações ou prescrever tratamentos para seus pacientes, em que grau você considera *diminuir a exposição do paciente a toxinas ambientais em:*

| | Sempre | Na maioria das vezes | Algumas vezes | Raramente | Nunca |
|---|---|---|---|---|---|
| Comida | ○ | ○ | ○ | ○ | ○ |
| Água | ○ | ○ | ○ | ○ | ○ |
| Ar | ○ | ○ | ○ | ○ | ○ |
| Produtos de cuidados pessoais (por exemplo, produtos de banho, cuidados com a pele, maquiagem) | ○ | ○ | ○ | ○ | ○ |

| | Sempre | Na maioria das vezes | Algumas vezes | Raramente | Nunca |
|---|---|---|---|---|---|
| Produtos para uso doméstico em ambientes internos (por exemplo, produtos de limpeza, materiais de construção) | ○ | ○ | ○ | ○ | ○ |
| Produtos para uso doméstico ao ar livre (por exemplo, jardinagem, controle de pragas) | ○ | ○ | ○ | ○ | ○ |
| Produtos, ferramentas e ambientes de trabalho | ○ | ○ | ○ | ○ | ○ |

### Block 3

**Clima e saúde ambiental na força de trabalho naturopática mais ampla**

As perguntas a seguir perguntam sobre seus pensamentos sobre o papel mais amplo dos naturopatas nas áreas de mudança climática e saúde ambiental.

Fora da prática clínica com pacientes individuais, alguns profissionais se envolvem em educação comunitária mais ampla sobre saúde por meio de **plataformas públicas, como workshops, palestras comunitárias, blogs/vlogs e mídias sociais**. Até que ponto você se envolve nesse tipo de educação pública sobre os seguintes tópicos de saúde?

| | Muitas vezes | Às vezes | Ocasionalmente | Raramente | Nunca |
|---|---|---|---|---|---|
| A conexão entre a saúde humana e o ambiente natural | ○ | ○ | ○ | ○ | ○ |
| Os impactos das mudança climáticas na saúde | ○ | ○ | ○ | ○ | ○ |
| Os impactos dos poluentes ambientais na saúde | ○ | ○ | ○ | ○ | ○ |
| Outros tópicos de saúde | ○ | ○ | ○ | ○ | ○ |

Até que ponto você acredita que, como naturopata/Médico Naturopata, você desempenha um papel no enfrentamento dos **impactos da mudança climática** na saúde de seus **pacientes individuais**?

- ○ Um papel muito significativo
- ○ Um papel significativo
- ○ Um papel um tanto significativo
- ○ Um papel ligeiramente significativo
- ○ Nenhuma função

Até que ponto você acredita que, como naturopata/Médico Naturopata, você desempenha um papel no enfrentamento dos **impactos da mudança climática** na sua **comunidade mais ampla**? (ou seja, além dos pacientes que você atende na clínica)

- Um papel muito significativo
- Um papel significativo
- Um papel um tanto significativo
- Um papel ligeiramente significativo
- Nenhuma função

Quão ***eficaz*** você acha que o tratamento naturopático é para lidar com o **impacto da mudança climática na saúde**?

- Muito eficaz
- Eficaz
- Um pouco eficaz
- Ligeiramente eficaz
- Não é nada eficaz

Quão ***confiante*** você se sente em fornecer cuidados naturopáticos para lidar com os **impactos da mudança climática na saúde**?

- Muito confiante
- Confiante
- Um pouco confiante
- Não muito confiante
- Nada confiante

Até que ponto você acha que precisa de **educação ou treinamento** adicional sobre o tema de como lidar com os **impactos da mudança climática na saúde**?

- Necessidade muito forte
- Forte necessidade
- Alguma necessidade
- Pequena necessidade
- Não preciso de nenhuma educação ou treinamento adicional

Até que ponto você acredita que, como naturopata/Médico Naturopata, você desempenha um papel no tratamento dos **impactos na saúde das toxinas ambientais** para seus **pacientes individuais**?

- Um papel muito significativo
- Um papel significativo
- Um papel um tanto significativo
- Um papel ligeiramente significativo
- Nenhuma função

Até que ponto você acredita que, como naturopata/Médico Naturopata, você desempenha um papel no enfrentamento dos **impactos na saúde das toxinas ambientais** para sua **comunidade mais ampla**? (ou seja, além dos pacientes que você atende na clínica)

- Um papel muito significativo
- Um papel significativo
- Um papel um tanto significativo
- Um papel ligeiramente significativo
- Nenhuma função

Quão ***eficaz*** você acha que o tratamento naturopático é para lidar com o **impacto na saúde das toxinas ambientais**?

- Muito eficaz
- Eficaz
- Um pouco eficaz
- Ligeiramente eficaz
- Não é nada eficaz

Quão ***confiante*** você se sente em fornecer cuidados naturopáticos para lidar com os **impactos na saúde dos poluentes tóxicos ambientais**?

- Muito confiante
- Confiante
- Um pouco confiante
- Um pouco confiante
- Nada confiante

Até que ponto você acha que precisa de ***educação ou treinamento adicional*** sobre o tópico **saúde ambiental e poluentes tóxicos**?

- Necessidade muito forte
- Forte necessidade
- Alguma necessidade
- Pequena necessidade
- Não preciso de nenhuma educação ou treinamento adicional

#### Block 4

#### Sobre você

*Para garantir que incluímos uma gama diversificada de perspectivas de praticantes naturopatas ao redor do mundo, forneça alguns detalhes sobre você abaixo. (Qualquer informação que você fornecer aqui permanecerá anônima)*

Em que país você mora?

Este é o mesmo país onde sua prática clínica está sediada?

- Sim
- Não (especifique onde você atua):

Qual das opções a seguir descreve melhor seu gênero?

- Feminino
- Masculino
- Não binário
- Prefiro me autodescrever:

Qual era sua idade no seu último aniversário?

Há quantos anos você atua como naturopata/médico naturopata?

- Até 10 anos
- 11 a 15 anos
- 16 a 20 anos
- 21 a 25 anos
- Mais de 25 anos

A sua prática tem um foco especializado? (selecione todas as opções aplicáveis)

- Não, minha prática clínica tem um ***foco geral*** sem especialização específica
- Sim, minha prática clínica tem foco especializado em **saúde ambiental**
- Sim, minha prática clínica tem foco especializado em **saúde relacionada ao clima**
- Sim, minha prática clínica tem foco especializado em **outro tópico** (especifique):

Qual opção descreve melhor o ambiente de prática clínica em seu **local de prática principal**?

- Estou sozinho em uma clínica
- Estou em uma clínica com outros profissionais de saúde, mas nenhum outro profissional naturopata
- Estou em uma clínica com outros profissionais naturopatas, mas nenhum outro tipo de profissional de saúde
- Estou em uma clínica com outros praticantes de naturopatia e outros profissionais de saúde
- Estou em um ambiente hospitalar
- Outra configuração

Qual das opções abaixo descreve melhor onde sua clínica está localizada?

- Área urbana ou suburbana
- Área regional (nem urbana nem rural)
- Área rural ou remota
- On-line

Powered by Qualtrics

Español

#### Info sheet

**Encuesta mundial de la profesión naturopática sobre el cambio climático y la salud ambiental[ETH24-10002]**

##### ¿QUIÉN ESTÁ LLEVANDO A CABO ESTA INVESTIGACIÓN?

Este estudio es una colaboración entre la Federación Mundial de Naturopatía (WNF) y el Consorcio Australiano de Investigación en Medicina Complementaria e Integrativa de la Universidad de Tecnología de Sídney (UTS:ARCCIM).

Mi nombre es Dra. Hope Foley y soy investigadora en UTS:ARCCIM y copresidenta del Comité de Salud Ambiental de la WNF. Estoy llevando a cabo esta investigación con la Dra. Iva Lloyd, directora ejecutiva de la WNF, y los miembros del equipo de investigación de UTS:ARCCIM, la profesora adjunta Amie Steel, el profesor adjunto Jon Adams, la Dra. Kirsten Baker y el Sr. Tristan Carter.

##### ¿DE QUÉ SE TRATA LA INVESTIGACIÓN?

El objetivo de esta investigación es comprender mejor las

perspectivas y prácticas de los profesionales de la naturopatía a nivel mundial con respecto a los impactos en la salud asociados con problemas ambientales como el cambio climático y los contaminantes ambientales.

Le invitamos a participar porque es un profesional de la naturopatía que pertenece a una asociación u organización profesional que es miembro de la WNF (la asociación u organización que se puso en contacto con usted en relación con esta investigación).

Antes de decidir participar en este estudio de investigación, por favor consulte los criterios de selección. Para participar en este estudio, usted debe:

- Haber estado activo en práctica clínica durante al menos 5 años
- Ser capaz de completar una encuesta en uno de los idiomas disponibles (inglés, español, portugués o francés)

**FINANCIACIÓN**
Este proyecto no ha recibido ninguna financiación.

**¿QUÉ IMPLICA MI PARTICIPACIÓN?**
Su participación en este estudio es voluntaria. Depende completamente de usted decidir si desea participar o no. Si decide participar, lo invitaremos a completar una

encuesta anónima en línea que le llevará aproximadamente 12 a 18 minutos.

Puede cambiar de opinión en cualquier momento y dejar de completar la(s) encuesta(s) sin consecuencias. Si decide no participar o no completar la encuesta, esto no afectará su relación con los investigadores, la UTS, la WNF o la asociación/organización que lo invitó. Su participación es anónima, por lo que nadie sabrá si ha participado o no.

**¿EXISTEN RIESGOS/INCONVENIENTES?**
Sí, existen algunos riesgos/inconvenientes. El tiempo que lleva completar la encuesta puede ser un inconveniente para algunas personas. Algunas personas también pueden tener el inconveniente de problemas de conectividad a Internet al intentar completar el cuestionario. Además, la encuesta incluye preguntas sobre el cambio climático y los contaminantes ambientales, temas que pueden resultar incómodos para algunas personas.

Hemos minimizado estos riesgos manteniendo la encuesta lo más breve posible y asegurándonos de que funcione en varios tipos de dispositivos. Las preguntas sobre el cambio climático y los contaminantes ambientales no son de naturaleza personal, sino que están relacionadas con sus perspectivas generales y prácticas profesionales, para reducir cualquier incomodidad, y no se le solicitará que proporcione ninguna información que no desee compartir. Si siente incomodidad o angustia, comuníquese con una persona de apoyo adecuada o un profesional de la salud.

**¿QUÉ PASARÁ CON MI INFORMACIÓN PERSONAL?**

Se accede al cuestionario a través del botón que se encuentra al final de esta página de información. Completar la encuesta es una indicación de su consentimiento. Como la encuesta es anónima, sus datos no pueden ser identificados ni eliminados después de completarla.

De acuerdo con las leyes de privacidad pertinentes de Australia y/o Nueva Gales del Sur, usted tiene derecho a solicitar acceso a la información sobre usted que recopila y almacena el equipo de investigación. También tiene derecho a solicitar que se corrija cualquier información con la que no esté de acuerdo. Informe al miembro del equipo de investigación nombrado al final de este documento si desea acceder a su información. Esto solo será posible si proporciona información personal que pueda identificarle individualmente o que sea razonablemente identificable (por ejemplo, si alguna respuesta de texto abierto le identifica o le vuelve a identificar contextualmente).

Se prevé que los resultados de este proyecto de

investigación serán publicados y/o presentados en una variedad de foros, incluidas las publicaciones y presentaciones de la WNF, y se puedan usar en proyectos futuros realizados por la WNF. Los datos de la investigación se almacenarán en una ubicación segura en las plataformas basadas en la nube de UTS (por ejemplo, OneDrive) a las que solo podrá acceder el equipo de investigación durante al menos 5 años. Los resultados de esta investigación también podrán ser compartidos a través de bases de datos científicos de acceso abierto (públicas), incluidas las bases de datos de Internet. Esto permitirá que otros investigadores utilicen los datos para investigar otras preguntas de investigación importantes. Los resultados compartidos de esta manera siempre se desidentificarán eliminando toda la información personal (si proporciona dicha información, por ejemplo, su nombre, dirección, fecha de nacimiento, etc.) y/o cualquier información contextual que pueda identificarle.

**¿QUÉ PASA SI TENGO ALGUNA DUDA O INQUIETUD?**
Si tiene alguna consulta o inquietud sobre la investigación con la que cree que podemos ayudar, no dude en comunicarse conmigo a. Alternativamente, si desea hablar con alguien de la WNF, puede comunicarse con la Dra. Iva Lloyd a

Si desea hablar con alguien que no esté relacionado con

la investigación, o si tiene alguna inquietud o queja sobre algún aspecto de la realización de esta investigación que desee plantear de forma independiente al equipo de investigación, comuníquese con la Secretaría de Ética al +61 2 9514 2478 o envíe un correo electrónico a y cite el número de referencia ETH24-10002 del UTS HREC. Cualquier asunto planteado será tratado de forma confidencial, se investigará y se le informará del resultado.

**CONSENTIMIENTO**
Antes de decidir participar en este estudio de investigación, verifique nuevamente los criterios de selección. Para participar en este estudio, usted debe:

- Haber estado activo en práctica clínica durante al menos 5 años
- Poder completar una encuesta en uno de los idiomas disponibles (inglés, español, portugués o francés)

**Encuesta mundial de la profesión naturopática sobre el cambio climático y la salud ambiental[ETH24-10002]**

**¿QUIÉN ESTÁ LLEVANDO A CABO ESTA INVESTIGACIÓN?**
Este estudio es una colaboración entre la Federación

Mundial de Naturopatía (WNF) y el Consorcio Australiano de Investigación en Medicina Complementaria e Integrativa de la Universidad de Tecnología de Sídney (UTS:ARCCIM).

Mi nombre es Dra. Hope Foley y soy investigadora en UTS:ARCCIM y copresidenta del Comité de Salud Ambiental de la WNF. Estoy llevando a cabo esta investigación con la Dra. Iva Lloyd, directora ejecutiva de la WNF, y los miembros del equipo de investigación de UTS:ARCCIM, la profesora adjunta Amie Steel, el profesor adjunto Jon Adams, la Dra. Kirsten Baker y el Sr. Tristan Carter.

**¿DE QUÉ SE TRATA LA INVESTIGACIÓN?**
El objetivo de esta investigación es comprender mejor las perspectivas y prácticas de los profesionales de la naturopatía a nivel mundial con respecto a los impactos en la salud asociados con problemas ambientales como el cambio climático y los contaminantes ambientales.

Le invitamos a participar porque es un profesional de la naturopatía que pertenece a una asociación u organización profesional que es miembro de la WNF (la asociación u organización que se puso en contacto con usted en relación con esta investigación).

Antes de decidir participar en este estudio de

investigación, por favor consulte los criterios de selección. Para participar en este estudio, usted debe:

- Haber estado activo en práctica clínica durante al menos 5 años
- Ser capaz de completar una encuesta en uno de los idiomas disponibles (inglés, español, portugués o francés)

**FINANCIACIÓN**

Este proyecto no ha recibido ninguna financiación.

**¿QUÉ IMPLICA MI PARTICIPACIÓN?**

Su participación en este estudio es voluntaria. Depende completamente de usted decidir si desea participar o no. Si decide participar, lo invitaremos a completar una encuesta anónima en línea que le llevará aproximadamente 12 a 18 minutos.

Puede cambiar de opinión en cualquier momento y dejar de completar la(s) encuesta(s) sin consecuencias. Si decide no participar o no completar la encuesta, esto no afectará su relación con los investigadores, la UTS, la WNF o la asociación/organización que lo invitó. Su participación es anónima, por lo que nadie sabrá si ha participado o no.

**¿EXISTEN RIESGOS/INCONVENIENTES?**

Sí, existen algunos riesgos/inconvenientes. El tiempo que lleva completar la encuesta puede ser un inconveniente para algunas personas. Algunas personas también pueden tener el inconveniente de problemas de conectividad a Internet al intentar completar el cuestionario. Además, la encuesta incluye preguntas sobre el cambio climático y los contaminantes ambientales, temas que pueden resultar incómodos para algunas personas.

Hemos minimizado estos riesgos manteniendo la encuesta lo más breve posible y asegurándonos de que funcione en varios tipos de dispositivos. Las preguntas sobre el cambio climático y los contaminantes ambientales no son de naturaleza personal, sino que están relacionadas con sus perspectivas generales y prácticas profesionales, para reducir cualquier incomodidad, y no se le solicitará que proporcione ninguna información que no desee compartir. Si siente incomodidad o angustia, comuníquese con una persona de apoyo adecuada o un profesional de la salud.

**¿QUÉ PASARÁ CON MI INFORMACIÓN PERSONAL?**
Se accede al cuestionario a través del botón que se encuentra al final de esta página de información. Completar la encuesta es una indicación de su consentimiento. Como la encuesta es anónima, sus datos no pueden ser identificados ni eliminados después

de completarla.

De acuerdo con las leyes de privacidad pertinentes de Australia y/o Nueva Gales del Sur, usted tiene derecho a solicitar acceso a la información sobre usted que recopila y almacena el equipo de investigación. También tiene derecho a solicitar que se corrija cualquier información con la que no esté de acuerdo. Informe al miembro del equipo de investigación nombrado al final de este documento si desea acceder a su información. Esto solo será posible si proporciona información personal que pueda identificarle individualmente o que sea razonablemente identificable (por ejemplo, si alguna respuesta de texto abierto le identifica o le vuelve a identificar contextualmente).

Se prevé que los resultados de este proyecto de investigación serán publicados y/o presentados en una variedad de foros, incluidas las publicaciones y presentaciones de la WNF, y se puedan usar en proyectos futuros realizados por la WNF. Los datos de la investigación se almacenarán en una ubicación segura en las plataformas basadas en la nube de UTS (por ejemplo, OneDrive) a las que solo podrá acceder el equipo de investigación durante al menos 5 años. Los resultados de esta investigación también podrán ser compartidos a través de bases de datos científicos de acceso abierto (públicas), incluidas las bases de datos de Internet. Esto permitirá que otros investigadores utilicen los datos para investigar otras preguntas de investigación importantes. Los resultados compartidos de esta manera siempre se desidentificarán eliminando toda la información personal (si proporciona dicha información, por ejemplo, su nombre, dirección, fecha de nacimiento, etc.) y/o cualquier información contextual que pueda identificarle.

#### ¿QUÉ PASA SI TENGO ALGUNA DUDA O INQUIETUD?

Si tiene alguna consulta o inquietud sobre la investigación con la que cree que podemos ayudar, no dude en comunicarse conmigo a. Alternativamente, si desea hablar con alguien de la WNF, puede comunicarse con la Dra. Iva Lloyd a

Si desea hablar con alguien que no esté relacionado con la investigación, o si tiene alguna inquietud o queja sobre algún aspecto de la realización de esta investigación que desee plantear de forma independiente al equipo de investigación, comuníquese con la Secretaría de Ética al +61 2 9514 2478 o envíe un correo electrónico a y cite el número de referencia ETH24-10002 del UTS HREC. Cualquier asunto planteado será tratado de forma confidencial, se investigará y se le informará del resultado.

#### CONSENTIMIENTO

Antes de decidir participar en este estudio de

investigación, verifique nuevamente los criterios de selección. Para participar en este estudio, usted debe:

- Haber estado activo en práctica clínica durante al menos 5 años
- Poder completar una encuesta en uno de los idiomas disponibles (inglés, español, portugués o francés)

○ Continuar con la encuesta
○ No deseo participar en la encuesta

#### Default Question Block

La siguiente encuesta le pregunta sobre sus perspectivas y experiencias en relación con aspectos del cambio climático y la salud ambiental en la práctica naturopática. Esta encuesta le llevará aproximadamente 15 minutos y sus respuestas son completamente anónimas.

#### Percepciones sobre el clima y la salud ambiental en la comunidad naturopática

Las preguntas de esta sección se refieren a sus perspectivas generales sobre los temas del cambio climático y la salud ambiental. Nos interesa saber qué piensan los médicos naturópatas sobre estos temas.

¿En qué medida está de acuerdo o en desacuerdo con las siguientes afirmaciones?

| | Estoy totalmente de acuerdo | Estoy de acuerdo | Ni de acuerdo ni en desacuerdo | Estoy en desacuerdo | Estoy en total desacuerdo |
|---|---|---|---|---|---|
| Creo que el cambio climático es real | ○ | ○ | ○ | ○ | ○ |

| | Estoy totalmente de acuerdo | Estoy de acuerdo | Ni de acuerdo ni en desacuerdo | Estoy en desacuerdo | Estoy en total desacuerdo |
|---|---|---|---|---|---|
| Las principales causas del cambio climático son las actividades humanas | ○ | ○ | ○ | ○ | ○ |
| El cambio climático traerá graves consecuencias negativas | ○ | ○ | ○ | ○ | ○ |
| Mi área local se verá afectada por el cambio climático | ○ | ○ | ○ | ○ | ○ |

| | Estoy totalmente de acuerdo | Estoy de acuerdo | Ni de acuerdo ni en desacuerdo | Estoy en desacuerdo | Estoy en total desacuerdo |
|---|---|---|---|---|---|
| Pasará mucho tiempo antes de que se sientan las consecuencias del cambio climático | ○ | ○ | ○ | ○ | ○ |
| El cambio climático puede afectar la salud humana | ○ | ○ | ○ | ○ | ○ |
| El cambio climático ya está afectando la salud humana | ○ | ○ | ○ | ○ | ○ |

#### ¿Qué tan de acuerdo o en desacuerdo está usted con las siguientes afirmaciones?

| | Estoy totalmente de acuerdo | Estoy de acuerdo | Ni de acuerdo ni en desacuerdo | Estoy en desacuerdo | Estoy en total desacuerdo |
|---|---|---|---|---|---|
| Me preocupan los posibles impactos del cambio climático en la salud humana. | ○ | ○ | ○ | ○ | ○ |
| Me preocupan los posibles impactos del cambio climático en la salud de mis pacientes. | ○ | ○ | ○ | ○ | ○ |

#### ¿Qué tan de acuerdo o en desacuerdo está usted con las siguientes afirmaciones?

| | Estoy totalmente de acuerdo | Estoy de acuerdo | Ni de acuerdo ni en desacuerdo | Estoy en desacuerdo | Estoy en total desacuerdo |
|---|---|---|---|---|---|
| Los contaminantes ambientales como los que se encuentran en el aire, el agua, los alimentos o los entornos domésticos están teniendo efectos negativos en la salud humana. | ○ | ○ | ○ | ○ | ○ |
| Me preocupan los efectos de los contaminantes ambientales en la salud humana. | ○ | ○ | ○ | ○ | ○ |
| Me preocupan los efectos de los contaminantes ambientales en la salud de mis pacientes. | ○ | ○ | ○ | ○ | ○ |

#### ¿Con qué frecuencia sus pacientes expresan las siguientes preocupaciones sobre el clima y la salud ambiental?

| | Siempre | La mayor parte del tiempo | A veces | Casi nunca | Nunca |
|---|---|---|---|---|---|
| Mis pacientes expresan preocupación por el impacto del cambio climático en su salud física | ○ | ○ | ○ | ○ | ○ |
| Mis pacientes expresan preocupación por el impacto del cambio climático en su salud mental | ○ | ○ | ○ | ○ | ○ |
| Mis pacientes expresan inquietudes sobre los contaminantes ambientales u otros factores ambientales que afectan su salud. | ○ | ○ | ○ | ○ | ○ |

#### Block 1

#### Salud climática y ambiental en la evaluación naturopática

La siguiente sección incluye preguntas sobre sus experiencias y perspectivas con respecto a la salud climática y ambiental en el contexto de la evaluación clínica naturopática.

Durante los últimos 5 años, ¿ha notado un aumento en la frecuencia con la que ve los siguientes síntomas y afecciones en su práctica clínica?

***Sí, he notado un aumento en... (seleccione todas las opciones que correspondan)***

- ☐ Asma, EPOC y/u otras afecciones respiratorias
- ☐ Cáncer
- ☐ Condiciones de deshidratación y/o golpe de calor
- ☐ Nefropatía
- ☐ Ansiedad, depresión y/u otros problemas de salud mental.
- ☐ Afecciones neurológicas
- ☐ Preocupaciones por la desnutrición
- ☐ Inseguridad alimentaria (bajo acceso a los alimentos)
- ☐ Enfermedades transmitidas por mosquitos
- ☐ Infertilidad femenina y disfunción reproductiva
- ☐ Infertilidad masculina y disfunción reproductiva
- ☐ Preocupaciones sobre el desarrollo infantil
- ☐ Diabetes
- ☐ Enfermedades autoinmunes
- ☐ Ninguna de las anteriores

En su opinión, ¿cuál de los siguientes factores están contribuyendo a este cambio/aumento?

| | Cambio climático | Contaminantes ambientales | Factores económicos | Factores sociales | Otro | No sé |
|---|---|---|---|---|---|---|
| » Asma, EPOC y/u otras afecciones respiratorias | ☐ | ☐ | ☐ | ☐ | ☐ | ☐ |
| » Cáncer | ☐ | ☐ | ☐ | ☐ | ☐ | ☐ |
| » Condiciones de deshidratación y/o golpe de calor | ☐ | ☐ | ☐ | ☐ | ☐ | ☐ |
| » Nefropatía | ☐ | ☐ | ☐ | ☐ | ☐ | ☐ |
| » Ansiedad, depresión y/u otros problemas de salud mental. | ☐ | ☐ | ☐ | ☐ | ☐ | ☐ |
| » Afecciones neurológicas | ☐ | ☐ | ☐ | ☐ | ☐ | ☐ |
| » Preocupaciones por la desnutrición | ☐ | ☐ | ☐ | ☐ | ☐ | ☐ |
| » Inseguridad alimentaria (bajo acceso a los alimentos) | ☐ | ☐ | ☐ | ☐ | ☐ | ☐ |
| » Enfermedades transmitidas por mosquitos | ☐ | ☐ | ☐ | ☐ | ☐ | ☐ |
| » Infertilidad femenina y disfunción reproductiva | ☐ | ☐ | ☐ | ☐ | ☐ | ☐ |

| | Cambio climático | Contaminantes ambientales | Factores económicos | Factores sociales | Otro | No sé |
|---|---|---|---|---|---|---|
| » Infertilidad masculina y disfunción reproductiva | ☐ | ☐ | ☐ | ☐ | ☐ | ☐ |
| » Preocupaciones sobre el desarrollo infantil | ☐ | ☐ | ☐ | ☐ | ☐ | ☐ |
| » Diabetes | ☐ | ☐ | ☐ | ☐ | ☐ | ☐ |
| » Enfermedades autoinmunes | ☐ | ☐ | ☐ | ☐ | ☐ | ☐ |
| » Ninguna de las anteriores | ☐ | ☐ | ☐ | ☐ | ☐ | ☐ |

¿En qué medida tiene en cuenta los siguientes factores relacionados con el clima durante la evaluación del caso de un paciente?

#### Alimentos y agua:

| | Siempre | La mayor parte del tiempo | Algunas veces | Casi nunca | Nunca |
|---|---|---|---|---|---|
| La **capacidad** del paciente para acceder a alimentos frescos y saludables. | ○ | ○ | ○ | ○ | ○ |

| | Siempre | La mayor parte del tiempo | Algunas veces | Casi nunca | Nunca |
|---|---|---|---|---|---|
| La sostenibilidad ambiental de los hábitos alimentarios de los pacientes (por ejemplo, abastecimiento local, baja huella de carbono) | ○ | ○ | ○ | ○ | ○ |
| La **capacidad** del paciente para acceder a agua potable limpia | ○ | ○ | ○ | ○ | ○ |
| Consumo de agua suficiente para el clima local y el estado de salud del paciente. | ○ | ○ | ○ | ○ | ○ |

### Calidad del aire y clima:

| | Siempre | La mayor parte del tiempo | Algunas veces | Casi nunca | Nunca |
|---|---|---|---|---|---|
| La exposición del paciente a la contaminación del aire externo debido al cambio climático o a fenómenos meteorológicos extremos (por ejemplo, humo de incendios forestales). | ○ | ○ | ○ | ○ | ○ |

| | Siempre | La mayor parte del tiempo | Algunas veces | Casi nunca | Nunca |
|---|---|---|---|---|---|
| El impacto de los cambios climáticos o fenómenos meteorológicos severos en la salud y el bienestar del paciente. | ○ | ○ | ○ | ○ | ○ |

#### ¿En qué medida tiene en cuenta los siguientes factores ambientales durante la evaluación del caso de un paciente?

**Alimentos y agua:**

| | Siempre | La mayor parte del tiempo | Algunas veces | Casi nunca | Nunca |
|---|---|---|---|---|---|
| Factores ambientales externos que afectan la calidad de los alimentos para los pacientes (por ejemplo, alimentos cultivados orgánicamente, evitar pesticidas o transgénicos) | ○ | ○ | ○ | ○ | ○ |

| | Siempre | La mayor parte del tiempo | Algunas veces | Casi nunca | Nunca |
|---|---|---|---|---|---|
| Factores ambientales domésticos que afectan la calidad de los alimentos para los pacientes (por ejemplo, toxinas de los recipientes para almacenar alimentos) | ○ | ○ | ○ | ○ | ○ |
| Factores ambientales externos que afectan la calidad del agua del paciente (por ejemplo, contaminación de fuentes de agua locales) | ○ | ○ | ○ | ○ | ○ |
| Factores ambientales domésticos que afectan la calidad del agua del paciente (por ejemplo, filtración y almacenamiento) | ○ | ○ | ○ | ○ | ○ |

#### Calidad del aire y ambiente general:

| | Siempre | La mayor parte del tiempo | Algunas veces | Casi nunca | Nunca |
|---|---|---|---|---|---|
| Exposición del paciente a **fuentes externas de contaminación del aire** en entornos de vida o trabajo (por ejemplo, gases de escape de automóviles, emisiones industriales) | ○ | ○ | ○ | ○ | ○ |
| Exposición del paciente a **fuentes domésticas/de interiores de contaminación del aire** en entornos de vida/trabajo (por ejemplo, productos de limpieza y cuidado personal, humo de tabaco, aparatos de cocina a gas y leña) | ○ | ○ | ○ | ○ | ○ |
| Exposición del paciente a **otras toxinas ambientales en el hogar, el trabajo o entornos naturales** (por ejemplo, frecuencias electromagnéticas, contaminación lumínica) | ○ | ○ | ○ | ○ | ○ |

| | Siempre | La mayor parte del tiempo | Algunas veces | Casi nunca | Nunca |
|---|---|---|---|---|---|
| **Realizar pruebas al paciente** para detectar la presencia de contaminantes ambientales tóxicos o impactos asociados en la salud (por ejemplo, pruebas de metales pesados y contaminantes orgánicos persistentes) | ○ | ○ | ○ | ○ | ○ |

#### Block 2

### Tratamientos y cuidados naturopáticos para la salud climática y ambiental

*Las preguntas de esta sección se refieren a sus experiencias y perspectivas sobre los tipos de cuidados, prescripciones y tratamientos utilizados por los naturópatas en relación con la salud climática y ambiental*

¿En qué medida incluye las siguientes opciones/factores al ***hacer recomendaciones o prescribir tratamientos*** a sus pacientes?

| | Siempre | La mayor parte del tiempo | Algunas veces | Casi nunca | Nunca |
|---|---|---|---|---|---|
| Recomendaciones basadas en alimentos y dietas ambientalmente sostenibles (por ejemplo, de origen local, de temporada, suministros abundantes, huella de carbono baja) | ○ | ○ | ○ | ○ | ○ |
| **Hierbas, suplementos y otros remedios recetados ambientalmente sostenibles (por ejemplo, de origen local, de temporada, en abundancia y con baja huella de carbono)** | ○ | ○ | ○ | ○ | ○ |
| La **calidad de los alimentos** que se recomiendan o consumen a los pacientes (por ejemplo, orgánicos, libres de pesticidas, criados en granjas versus capturados en la naturaleza, a base de plantas) | ○ | ○ | ○ | ○ | ○ |
| Aumentar el tiempo que los pacientes pasan en la naturaleza (prescripción de naturaleza) | ○ | ○ | ○ | ○ | ○ |

| | Siempre | La mayor parte del tiempo | Algunas veces | Casi nunca | Nunca |
|---|---|---|---|---|---|
| Educar a los pacientes sobre la **conexión entre la salud humana y el medio ambiente natural** | ○ | ○ | ○ | ○ | ○ |
| Educar a los pacientes sobre los **impactos de los contaminantes ambientales sobre la salud** | ○ | ○ | ○ | ○ | ○ |

Al hacer recomendaciones o prescribir tratamientos a sus pacientes, ¿en qué medida considera *disminuir la exposición del paciente a toxinas ambientales en:*

| | Siempre | La mayor parte del tiempo | Algunas veces | Casi nunca | Nunca |
|---|---|---|---|---|---|
| Alimento | ○ | ○ | ○ | ○ | ○ |
| Agua | ○ | ○ | ○ | ○ | ○ |
| Aire | ○ | ○ | ○ | ○ | ○ |
| Productos de cuidado personal (por ejemplo, productos de baño, cuidado de la piel, maquillaje) | ○ | ○ | ○ | ○ | ○ |

| | Siempre | La mayor parte del tiempo | Algunas veces | Casi nunca | Nunca |
|---|---|---|---|---|---|
| Productos de uso doméstico en interiores (por ejemplo, productos de limpieza, materiales de construcción) | ○ | ○ | ○ | ○ | ○ |
| Productos para uso doméstico en exteriores (por ejemplo, jardinería, control de plagas) | ○ | ○ | ○ | ○ | ○ |
| Productos, herramientas y entornos de trabajo | ○ | ○ | ○ | ○ | ○ |

#### Block 3

**Salud climática y ambiental en el ámbito laboral naturopático en general**

Las siguientes preguntas se refieren a sus opiniones sobre el rol más amplio de los naturópatas en los campos del cambio climático y la salud ambiental.

Además de la práctica clínica con pacientes individuales, algunos profesionales participan en una educación comunitaria más amplia sobre la salud a través de **plataformas públicas como talleres, charlas comunitarias, blogs/vlogs y redes sociales**. ¿En qué medida participa en este tipo de educación pública sobre los siguientes temas de salud?

| | Frecuentemente | A veces | Ocasionalmente | Casi nunca | Nunca |
|---|---|---|---|---|---|
| La conexión entre la salud humana y el medio ambiente natural | ○ | ○ | ○ | ○ | ○ |
| Los efectos del cambio climático sobre la salud | ○ | ○ | ○ | ○ | ○ |
| Los efectos de los contaminantes ambientales sobre la salud | ○ | ○ | ○ | ○ | ○ |
| Otros temas de salud | ○ | ○ | ○ | ○ | ○ |

¿En qué medida cree que usted, como naturópata/ND, desempeña un papel a la hora de abordar los **impactos del cambio climático** en la salud de sus **pacientes individuales**?

- ○ Un papel muy importante
- ○ Un papel importante
- ○ Un papel bastante significativo
- ○ Un papel ligeramente significativo
- ○ Ningún papel en absoluto

¿En qué medida cree que usted, como naturópata/ND, desempeña un papel en el abordar de los **impactos de salud del cambio climático** para su **comunidad en general**? (es decir, más allá de los pacientes que ve en la clínica)

- Un papel muy importante
- Un papel importante
- Un papel bastante significativo
- Un papel ligeramente significativo
- Ningún papel en absoluto

¿Qué tan ***efectiva*** cree usted que es la atención naturopática para abordar el **impacto del cambio climático en la salud**?

- Muy eficaz
- Eficaz
- Algo efectivo
- Ligeramente eficaz
- No es efectivo en absoluto

¿Qué tan ***seguro(a)*** se siente en brindar atención naturopática para abordar los **impactos del cambio climático en la salud**?

- Muy seguro(a)
- Seguro(a)
- Algo seguro(a)
- No muy seguro(a)
- No me siento seguro(a) en absoluto

¿En qué medida cree que necesita **educación o capacitación** adicional sobre el tema de cómo abordar los **impactos del cambio climático en la salud**?

- Necesidad muy fuerte
- Necesidad fuerte
- Nececidad moderada
- Ligera necesidad
- No necesito ninguna educación o formación adicional.

¿En qué medida cree que usted, como naturópata/ND, desempeña un papel a la hora de abordar los **impactos de las toxinas ambientales** en la salud de sus **pacientes individuales**?

- Un papel muy importante
- Un papel importante
- Un papel bastante significativo
- Un papel ligeramente significativo
- Ningún papel en absoluto

¿En qué medida cree que usted, como naturópata/ND, desempeña un papel en el abordaje de los **impactos de las toxinas ambientales** en la salud de su **comunidad en general**? (es decir, más allá de los pacientes que ve en la clínica)

- Un papel muy importante
- Un papel importante
- Un papel bastante significativo
- Un papel ligeramente significativo
- Ningún papel en absoluto

¿Qué tan ***efectiva*** cree usted que es la atención naturopática para abordar el **impacto de las toxinas ambientales en la salud**?

- Muy efectiva
- Efectiva
- Algo efectiva
- Ligeramente efectiva
- No es efectiva en absoluto

¿Qué tan ***seguro(a)*** se siente de brindar atención naturopática para abordar los **impactos en la salud de**

**los contaminantes ambientales tóxicos**?

- Muy seguro(a)
- Seguro(a)
- Algo seguro(a)
- Un poco seguro(a)
- No me siento seguro(a) en absoluto

¿En qué medida considera que necesita ***educación o formación adicional*** sobre el tema de la **salud ambiental y los contaminantes tóxicos**?

- Necesidad muy fuerte
- Necesidad fuerte
- Necesidad moderada
- Ligera necesidad
- No necesito ninguna educación o formación adicional.

#### Block 4

#### Acerca de usted

*Para asegurarnos de que hemos incluido una amplia gama de perspectivas de profesionales naturopáticos de todo el mundo, proporcione algunos detalles sobre usted a continuación. (Cualquier información que proporcione aquí permanecerá anónima)*

¿En qué país vive?

¿Es este el mismo país en el que se basa su práctica clínica?

- Sí
- No (especifique dónde ejerce):

¿Cuál de las siguientes opciones describe mejor su género?

- Femenino
- Masculino
- No binario
- Prefiero autodescribirme:

¿Qué edad tenía en su último cumpleaños?

¿Cuántos años lleva ejerciendo como médico naturópata/naturópata?

- ○ 10 años o menos
- ○ 11 a 15 años
- ○ 16 a 20 años
- ○ 21 a 25 años
- ○ Más de 25 años

¿Su práctica tiene un enfoque especializado? (seleccione todas las opciones que correspondan)

- ☐ No, mi práctica clínica tiene un ***enfoque general*** sin ninguna especialización particular.
- ☐ Sí, mi práctica clínica tiene un enfoque especializado en **salud ambiental**
- ☐ Sí, mi práctica clínica tiene un enfoque especializado en la **salud relacionada con el clima**.
- ☐ Sí, mi práctica clínica tiene un enfoque especializado en **otro tema** (especifique):

¿Qué opción describe mejor el entorno de práctica clínica en su **lugar de práctica principal**?

- Estoy solo/sola en una clínica
- Estoy en una clínica con otros profesionales de la salud pero sin otros profesionales naturópatas.
- Estoy en una clínica con otros profesionales naturópatas pero no con ningún otro tipo de profesional de la salud.
- Estoy en una clínica con otros médicos naturópatas y otros profesionales de la salud.
- Estoy en un entorno hospitalario.
- Otro entorno clínico

¿Cuál de las siguientes opciones describe mejor dónde se encuentra su práctica clínica?

- [ ] Área urbana o suburbana
- [ ] Área regional (ni urbana ni rural)
- [ ] Zona rural o remota
- [ ] En línea

Powered by Qualtrics
